# Integrating Single-Molecule Variant Phenotyping with Clinical Features Predicts Outcomes in KIF1A-Associated Neurological Disorder

**DOI:** 10.64898/2026.09.10.26362467

**Authors:** Lu Rao, Wenxing Li, Jessica Waxler, Candace Cameron, Celia Tam, Yichun Wang, Charles A. LeDuc, Yufeng Shen, Arne Gennerich, Wendy K. Chung

**Affiliations:** Department of Biochemistry and Gruss Lipper Biophotonics Center, Albert Einstein College of Medicine, Bronx, NY 10461; Department of Systems Biology, Columbia University Irving Medical Center, New York, NY 10032; Department of Pediatrics, Boston Children’s Hospital, Boston, MA 02115; Harvard Medical School, Boston, MA 02115; Department of Biomedical Informatics, Columbia University Irving Medical Center, New York, NY 10032; Department of Biological Science, Columbia University, New York, NY 10027

**Keywords:** KIF1A-associated neurological disorder, adaptive behavior trajectory, genotype-phenotype association, single-molecule motility, machine learning prediction

## Abstract

Rare monogenic diseases lack scalable prognostic frameworks, leaving patients and families without guidance after molecular diagnosis. Using KIF1A-associated neurological disorder (KAND) as a model system, we integrated single-molecule biophysical phenotyping of 91 pathogenic variants with longitudinal clinical characterization of 343 patients. Vineland Adaptive Behavior Scales (VABS) Adaptive Behavior Composite (ABC) and Growth Scales identified two divergent trajectories emerging after age 10: a stable group and a declining group characterized by seizures, abnormal EEGs, and optic nerve atrophy. A framework combining biophysical parameters, computational pathogenicity scores, and clinical variables classified patients as stable versus declining with AUC 0.834 and predicted VABS ABC scores with R² = 0.484 under leave-one-out cross-validation. For newly identified KIF1A variants, this approach offers a prognostic tool and a stratification strategy for clinical trials. Because kinesin motor function is mechanistically conserved, the framework may generalize to other kinesinopathies and to monogenic disorders amenable to quantitative functional assays.

## 1. Introduction

Rare monogenic diseases collectively affect tens of millions of individuals, yet a fundamental gap persists between molecular diagnosis and clinical prognosis. Once a pathogenic variant is identified, families and clinicians frequently face the same question: what does this variant mean for this individual’s future? Conventional approaches to answering this question rely on prior case observations or population-level genetic inferences, neither of which is possible when a variant is newly identified, ultra-rare, or absent from population databases—the rule rather than the exception in rare monogenic disease [1, 2]. Computational pathogenicity scores partially address this gap [3] but were designed to classify variants as pathogenic or benign, rather than to quantify disease severity along a continuous axis. Quantitative functional assays, by directly measuring the biophysical consequences of each variant, offer a complementary and scalable alternative that does not depend on prior clinical observations [3–7]. Whether such measurements can be integrated with longitudinal clinical data to generate individualized outcome predictions in a rare monogenic disorder has not been systematically demonstrated.

KIF1A-associated neurological disorder (KAND) exemplifies this challenge. KAND is a group of severe, progressive rare diseases caused by pathogenic variants in *KIF1A* [8], a gene encoding a neuron-specific kinesin-3 motor protein responsible for anterograde axonal transport of synaptic vesicle precursors, dense-core vesicles, and other neuronal cargoes [9, 10]. The essential role of *KIF1A* in axonal cargo delivery means that disruption of its motor function directly impairs synaptic vesicle transport, synaptogenesis, and neuronal viability [11, 12]. More than 500 individuals worldwide have been diagnosed, and KAND encompasses a wide clinical spectrum ranging from mild hereditary spastic paraplegia to developmental epileptic encephalopathy with intellectual disability, optic nerve atrophy, and cerebellar degeneration [3, 13]. The vast majority of cases are *de novo* heterozygous missense variants in the motor domain, which exert dominant-negative effects by impairing the motility of heterodimeric motors composed of mutant and wild-type KIF1A subunits [6, 14, 15]. Even individuals carrying the same variant can exhibit substantially different disease courses [16], indicating that variant identity alone is insufficient to predict outcomes and that integrating variant-and patient-level information is required.

The Vineland Adaptive Behavior Scales (VABS) Adaptive Behavior Composite (ABC) score has emerged as a scalable quantitative outcome measure in KAND, capturing changes in communication, daily living, and socialization across the lifespan [3, 13]. Prior work has established several important genotype–phenotype relationships: seizures and abnormal EEG findings are the strongest clinical predictors of VABS decline [13]; computational pathogenicity scores, particularly the ESM [17] and MisFit [18], associate with VABS outcomes [6, 13]; and single-molecule total internal reflection fluorescence (smTIRF) microscopy has demonstrated that disease-associated KIF1A variants produce distinct mechanochemical defects, including reduced velocity, shortened run length, and loss of processive motility, that correlate broadly with clinical severity [4, 6, 19]. Despite these advances, prior studies have examined molecular and clinical predictors largely in isolation, and no framework has integrated these complementary data layers to generate individualized longitudinal predictions.

The need for such a framework is heightened by the emergence of disease-modifying therapies for KAND, including antisense oligonucleotide strategies [20, 21]. Designing rigorous clinical trials for a disorder as heterogeneous as KAND requires accurate prediction of patient trajectories to enable appropriate patient selection and outcome monitoring [22].

Here we address this question through the largest systematic integration of single-molecule biophysics and quantitative clinical phenotyping performed to date for any molecular motor disease. We characterized the single-molecule motility properties of 91 KIF1A disease variants using smTIRF microscopy and correlated these biophysical parameters with deep clinical phenotyping of 343 individuals with pathogenic or likely pathogenic *KIF1A* variants, of whom 238 had longitudinal VABS assessments. We developed a machine-learning prediction model that integrates molecular, computational, and clinical data to enable severity stratification and VABS prediction in individuals with KAND, and tests whether single-molecule biophysical phenotyping can serve as a scalable approach to refine prognosis in rare monogenic disorders.

## 2. Results

### 2.1 KAND Patients Bifurcate into Two Adaptive Behavior Trajectories After Age 10

Among 313 individuals carrying pathogenic or likely pathogenic KIF1A variants, we identified 291 missense variants (110 unique) and 22 loss-of-function (LoF) variants. A total of 106 unique missense variants (96.4%) were localized within the kinesin motor domain (**Supplementary Fig. 1**). Five highly recurrent variants (R254W, n = 31; R316W, n = 29; T99M, n = 17; P305L, n = 12; and R307Q, n = 12) collectively accounted for 101 individuals (35%). To characterize the clinical spectrum of KAND, we analyzed longitudinal VABS assessments from 238 individuals with pathogenic or likely pathogenic KIF1A variants. The distribution of most recent VABS ABC scores was unimodal in patients aged ≤10 years but bimodal in those aged >10 years (**Fig. 1a**), suggesting two distinct clinical subgroups emerging over time. Trajectory-based clustering of patients with two or more longitudinal VABS assessments identified Group 1 (higher and stable scores, n = 97) and Group 2 (lower and declining scores, n = 54), with 87 patients unclassified due to insufficient longitudinal data (**Fig. 1b**). The two trajectory patterns increasingly diverged after age 10, with Group 2 patients showing progressive decline. This age-10 inflection point was supported by analysis of longitudinal VABS dynamics: both the change in VABS ABC and the rate of change were more variable before age 10 and stabilized thereafter (**Supplementary Fig. 2**), supporting the most recent VABS ABC score as the primary outcome measure for trajectory analysis.

**Fig. 1.**
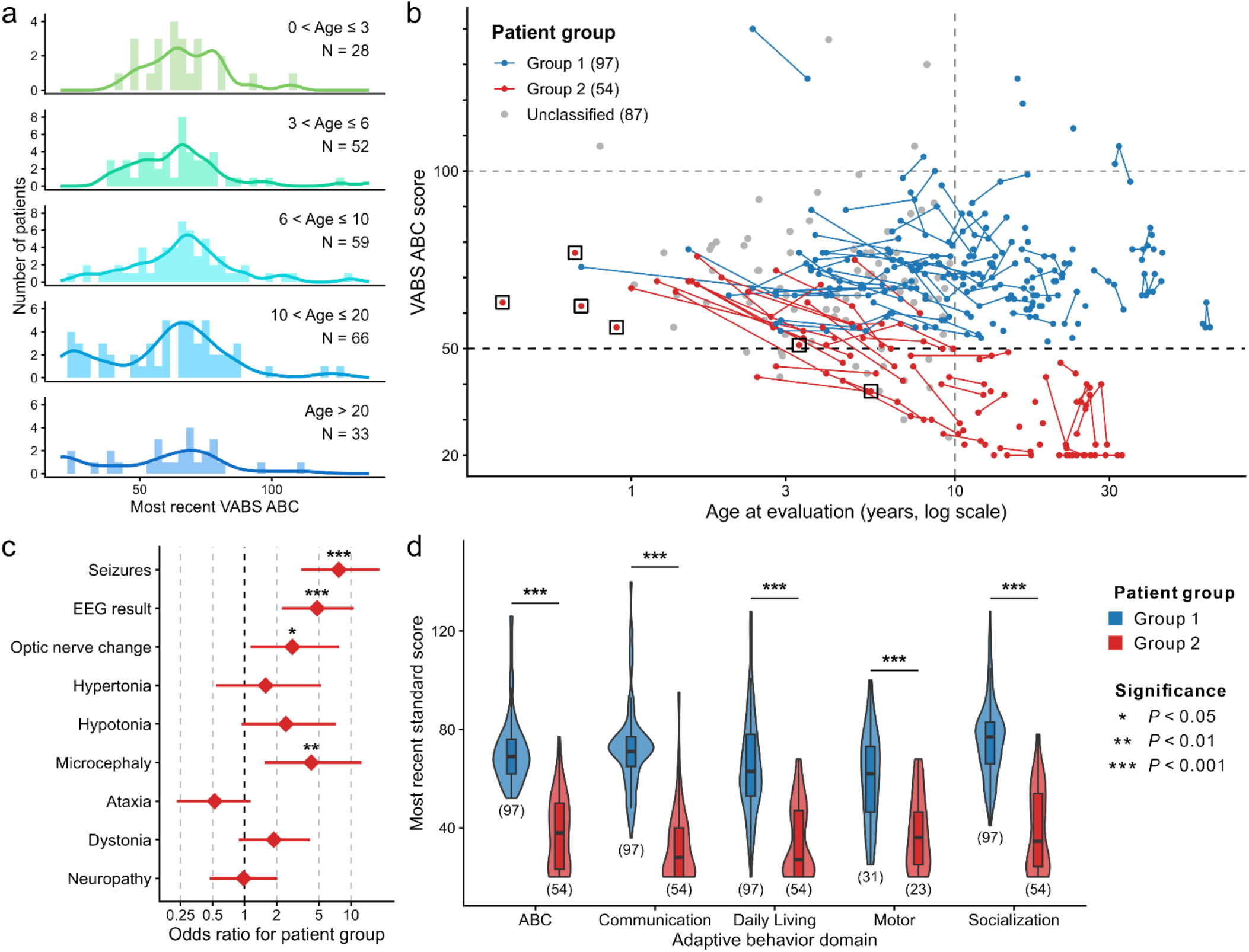
VABS ABC trajectories and clinical phenotypes in KAND patients. **(a)** Distribution of most recent VABS ABC scores across age groups. The distribution is unimodal in patients aged ≤ 10 years, while a bimodal pattern emerges in patients aged > 10 years. **(b)** Longitudinal VABS ABC trajectories for all patients in this study. Among patients with two or more VABS assessments, trajectories were used to classify individuals into two groups: Group 1 (higher and stable) and Group 2 (lower and rapid decline). Hollow squares indicate patients who died before age 10, who were assigned to Group 2. After age 10, two distinct trajectory patterns become apparent. **(c)** Differences in clinical phenotypes between patient groups. Compared to Group 1, Group 2 showed significantly higher odds of seizures, abnormal EEG results, optic nerve changes, microcephaly, and dystonia. Significance was assessed using Fisher’s exact test. **(d)** Differences in most recent VABS domain scores between patient groups. The numbers in parentheses indicate the sample size. Significance was assessed using Wilcoxon rank-sum test. Significance: * *P* < 0.05, ** *P* < 0.01, *** *P* < 0.001.

Compared with Group 1, Group 2 patients had significantly higher odds of seizures, abnormal EEGs, optic nerve changes, microcephaly, and dystonia (**Fig. 1c**, **Supplementary Table 1**) and scored significantly lower across all VABS adaptive behavior domains, including communication, daily living, motor, and socialization (**Fig. 1d**), confirming broader functional impairment. Unclassified patients more closely resembled Group 1 than Group 2 in VABS ABC scores, seizure frequency, and EEG abnormalities (**Supplementary Table 1**), suggesting enrichment for stable-trajectory patients who may emerge as Group 1-like with additional longitudinal follow-up.

To compare adaptive skill acquisition between Group 1 and Group 2, we examined growth scale values (GSV) across all 11 Vineland-3 subdomains. Unlike age-normed standard scores, GSVs provide an age-independent measure of skill level and increase monotonically in typical development, making them suitable for longitudinal within-person comparisons [23]. Group 1 patients continued with a positive GSV over the observation period, reflecting ongoing acquisition of new skills, whereas Group 2 patients showed median GSV rates of change approaching zero across most subdomains spanning communication, daily living skills, and socialization (**Supplementary Fig. 3**). Within Group 2, the trajectories were heterogeneous: most patients maintained lower absolute GSV values and slower rates of change than Group 1, while a subset remained at floor-level GSV with negligible change, indicating little to no acquisition of new skills (**Supplementary Fig. 4**).

### 2.2 Single-Molecule Motility Phenotyping Resolves Two Molecular Clusters linked to Patient Group

To investigate how individual variants affect KIF1A’s single-molecule behavior, we used an *E. coli* expression system to produce a tail-truncated, dimerized KIF1A construct containing the motor’s native neck coil followed by a leucine zipper, which recapitulates the behavior of full-length activated KIF1A [4, 5]. After purification and fluorescent labeling, key motility parameters including velocity, run length, motility status, and diffusional behavior were extracted by smTIRF microscopy (**Supplementary Fig. 5**). The breadth of phenotypic variation captured by this approach is illustrated by four structurally proximate variants within the switch-I loop: A206V retained near-wild-type motility, M210T exhibited reduced velocity, S215R displayed strong microtubule binding without processive movement, and R216C failed to bind microtubules altogether (**Supplementary Fig. 5**). This dissociation between structural location and functional consequence highlights the necessity of direct single-molecule characterization rather than inference from structural proximity alone.

Across 91 unique variants spanning 66 distinct amino acid positions, 22 positions harbored multiple substitutions. 33 positions (50%) correspond to residues that are highly conserved across kinesins (>90% similarity). Functionally, 38% of variants exhibited no detectable movement, 46% displayed diffusional behavior, and 26% showed prolonged microtubule dwell times relative to wild type (**Fig. 2a**). Among the 57 variants that did show directional movement, 95% moved at reduced velocities and all exhibited shorter run lengths than wild type, indicating broad motility impairment across variants (**Fig. 2a**).

**Fig. 2.**
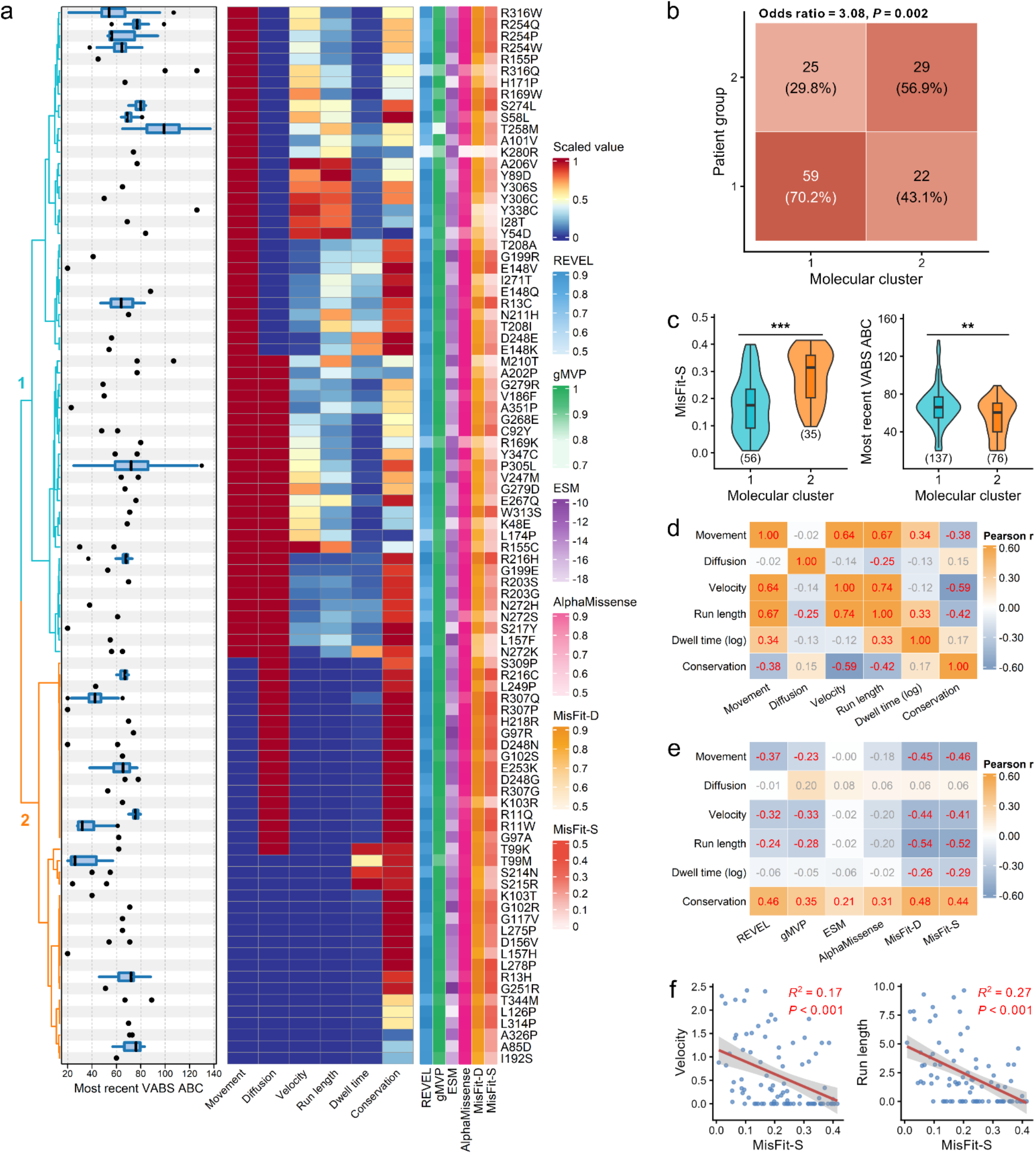
Single-molecule phenotypes of KIF1A variants. **(a)** Hierarchical clustering heatmap of single-molecule data for 91 KIF1A variants. The left panel shows the distribution of most recent VABS ABC scores for patients carrying each variant, and the right panel displays the corresponding computational pathogenicity scores. All molecular features were scaled to the range [0, 1] prior to clustering. Based on the hierarchical clustering results, variants were classified into two molecular clusters (cyan for Cluster 1, orange for Cluster 2). **(b)** Contingency table of molecular cluster and patient group assignments. Compared to Cluster 1, Cluster 2 shows a higher proportion of Group 2 patients (Fisher’s exact test). **(c)** Compared to Cluster 1, Cluster 2 exhibits significantly higher MisFit-S scores and lower most recent VABS ABC scores (Wilcoxon rank-sum test). The numbers in parentheses indicate the sample size. Significance: * *P* < 0.05, ** *P* < 0.01, *** *P* < 0.001. **(d)** Pairwise correlations among single-molecule features. Numbers in each cell indicate Pearson correlation coefficients; red values indicate statistically significant correlations (P < 0.05). **(e)** Pairwise correlations between single-molecule features and computational pathogenicity scores. **(f)** Representative examples showing that MisFit-S is significantly negatively correlated with both velocity and run length.

To complement these single-molecule measurements, we compiled six computational pathogenicity scores per variant: REVEL [24], gMVP [25], and AlphaMissense [26] to predict variant pathogenicity; ESM [17] to reflect evolutionary constraint; and MisFit-D and MisFit-S [18] to estimate molecular damage and the heterozygous selection coefficient, respectively. All scores except AlphaMissense were significantly correlated with the most recent VABS ABC (P < 0.05). However, the distributions of gMVP and AlphaMissense were heavily concentrated in the 0.9–1.0 range, limiting their resolving power across the variant spectrum (**Supplementary Fig. 6**).

Hierarchical clustering of the single-molecule motility properties of the 91 unique *KIF1A* variants (Methods) used scaled molecular features, including movement status (motile vs. non-motile), diffusion behavior (back-and-forth movement along microtubules), velocity, run length, dwell time, and conservation score (**Fig. 2a**). The clustering revealed two molecular clusters distinguished primarily by run length: Cluster 2 variants exhibited a complete loss of processive motility, whereas Cluster 1 variants retained varying degrees of motor activity (**Fig. 2a**). Compared with Cluster 1, Cluster 2 contained a significantly higher proportion of Group 2 patients (56.9% vs. 29.8%; odds ratio = 3.08, *P* = 0.002, Fisher’s exact test; **Fig. 2b**) and exhibited significantly higher MisFit-S scores and lower most recent VABS ABC scores (**Fig. 2c**), establishing a direct link between molecular cluster membership and clinical phenotypic group.

Pairwise correlation analysis revealed strong inter-correlations among single-molecule features, particularly between velocity and run length (r = 0.74) and between movement status and velocity (r = 0.64). Conservation was negatively correlated with movement-related features (**Fig. 2d**). Computational scores REVEL, gMVP, MisFit-D, and MisFit-S were each negatively correlated with movement status, velocity, and run length, while diffusion behavior was not significantly correlated with any pathogenicity score (**Fig. 2e**). MisFit-S was significantly negatively correlated with both velocity (r² = 0.17, *P* < 0.001) and run length (r² = 0.27, *P* < 0.001), with higher MisFit-S scores associated with lower velocity and shorter run length (**Fig. 2f**). These findings demonstrate that single-molecule motility phenotyping, particularly features related to microtubule interaction and processive motility, provides prognostic information that complements computational scoring.

### 2.3 Clinical, Computational, and Single-Molecule Features Are Each Associated with VABS Outcomes

To assess the prognostic information carried by different data layers, we examined the associations between adaptive behavior outcomes and three categories of features: clinical phenotypes, computational pathogenicity scores, and single-molecule motility parameters (**Fig. 3**, **Supplementary Tables 2, 3**).

**Fig. 3.**
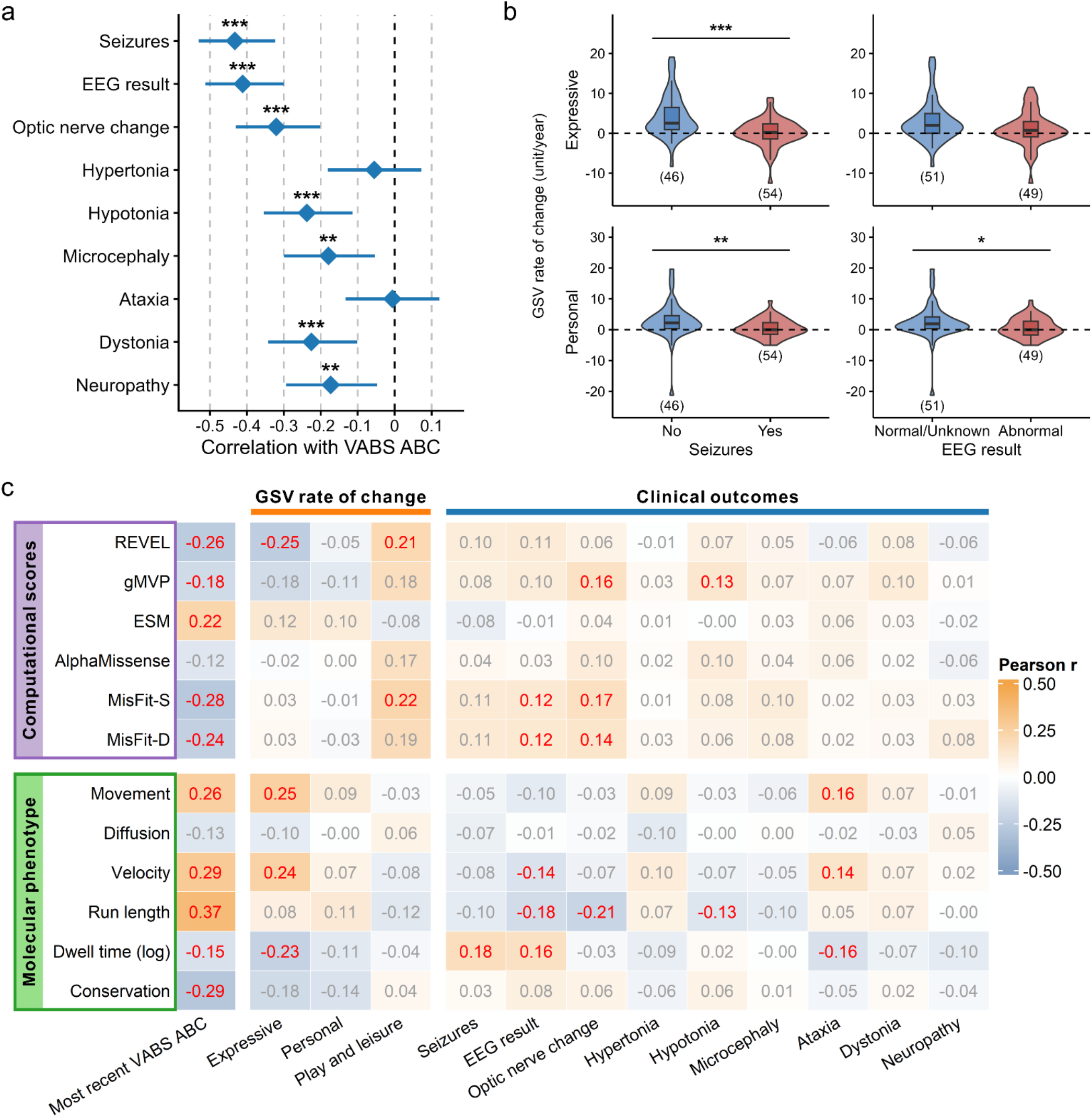
Associations among clinical outcomes, computational scores, and molecular features. **(a)** Clinical phenotypes significantly negatively correlated with most recent VABS ABC score (Pearson correlation). **(b)** Compared to patients without seizures or with normal/unknown EEG results, patients with seizures or abnormal EEG results showed significantly reduced growth scale value (GSV) rate of change in expressive communication and personal daily living skills (Wilcoxon rank-sum test). The numbers in parentheses indicate the sample size. Significance: * *P* < 0.05, ** *P* < 0.01, *** *P* < 0.001. **(c)** Heatmap showing Pearson correlations between computational scores or molecular phenotype features and clinical outcomes. Numbers in each cell indicate Pearson correlation coefficients; red values indicate statistically significant correlations (*P* < 0.05).

Clinical phenotypes, including seizures, abnormal EEG, optic nerve change, hypotonia, microcephaly, and dystonia were each significantly negatively correlated with the most recent VABS ABC (**Fig. 3a**, **Supplementary Table 2**). Patients with seizures or abnormal EEG showed significantly reduced GSV rates of change in expressive communication and personal daily living skills compared to individuals without seizure or abnormal EEG (**Fig. 3b, Supplementary Table 3**), indicating that these clinical features are associated with a reduced rate of adaptive skill acquisition.

Heatmap analysis of Pearson correlations between molecular and computational features and clinical outcomes revealed distinct patterns of association across feature types (**Fig. 3c**). Among computational scores, ESM was positively correlated with VABS ABC (r = 0.22), while MisFit-S and MisFit-D were negatively correlated (r = −0.28 and −0.24, respectively) and positively correlated with seizures and EEG abnormalities. The opposite signs reflect scoring conventions: less negative ESM scores indicate less evolutionary deviation, while higher MisFit-D and MisFit-S scores indicate greater molecular damage and stronger purifying selection, respectively, but all three converge on worse clinical outcomes for more deleterious variants. REVEL was negatively correlated with GSV rate of change in expressive communication (r = −0.25), further linking computational pathogenicity to impaired language skill acquisition. Nominal positive correlations between computational scores and GSV rate of change in the play and leisure subdomain were observed in the full cohort; however, sensitivity analysis demonstrated that these associations were driven by a single high-influence observation and were not significant upon its removal (both P > 0.05, **Supplementary Fig. 7**), and are therefore not interpreted as reflecting a true biological relationship.

Among single-molecule features (**Fig. 3c**), run length showed the strongest association with VABS ABC (r = 0.37), followed by velocity (r = 0.29) and movement status (r = 0.26). Dwell time and conservation were negatively correlated (r = −0.15 and −0.29). Run length was also negatively correlated with EEG abnormalities, optic nerve change, and hypotonia (r = −0.18, −0.21, and −0.13, respectively). Dwell time was positively correlated with seizures and abnormal EEG results (r = 0.18 and 0.16) and negatively correlated with ataxia (r = −0.16). Notably, run length was the strongest single-molecule correlate of VABS ABC, consistent with its central role in distinguishing the two molecular clusters. For GSV rate of change (**Fig. 3c**), movement status and velocity were positively correlated with expressive language (r = 0.25 and 0.24), and Dwell time was negatively correlated with play and leisure (r = −0.23).

Clinical features showed the strongest associations with VABS ABC, but multiple molecular and computational features were also significantly correlated, supporting the inclusion of all three feature types as predictors in the machine learning model.

### 2.4 An Integrated Two-Stage Model Predicts VABS ABC Score and Patient Group

To predict VABS ABC scores in individuals with KAND, we developed a two-stage machine learning framework that integrates molecular, computational, and clinical features (**Fig. 4a**). A key challenge in rare disease research is limited sample size, which requires maximizing the use of all available data while avoiding data leakage between training and test sets—a particular concern when patients share recurrent variants and therefore identical molecular features. To address this, we separated variant-level and patient-level modeling. Stage 1, a variant-level model, used computational pathogenicity scores and single-molecule motility features to predict the mean VABS ABC per variant and was evaluated using leave-one-variant-out cross-validation (LOVO-CV), which ensures that each variant is predicted solely from models trained on the remaining variants. Stage 2, a patient-level model, combined the Stage 1 variant-level predictions with patient-level clinical features (age at VABS assessment, EEG/seizure status, and additional clinical variables) to predict the most recent VABS ABC and patient group, and was also evaluated by LOVO-CV. Random Forest was selected as the base algorithm for both stages, providing a balance of predictive performance and interpretability. The model was applied to 214 patients with 79 unique variants for whom both VABS data and either molecular phenotype or computational scores were available.

**Fig. 4.**
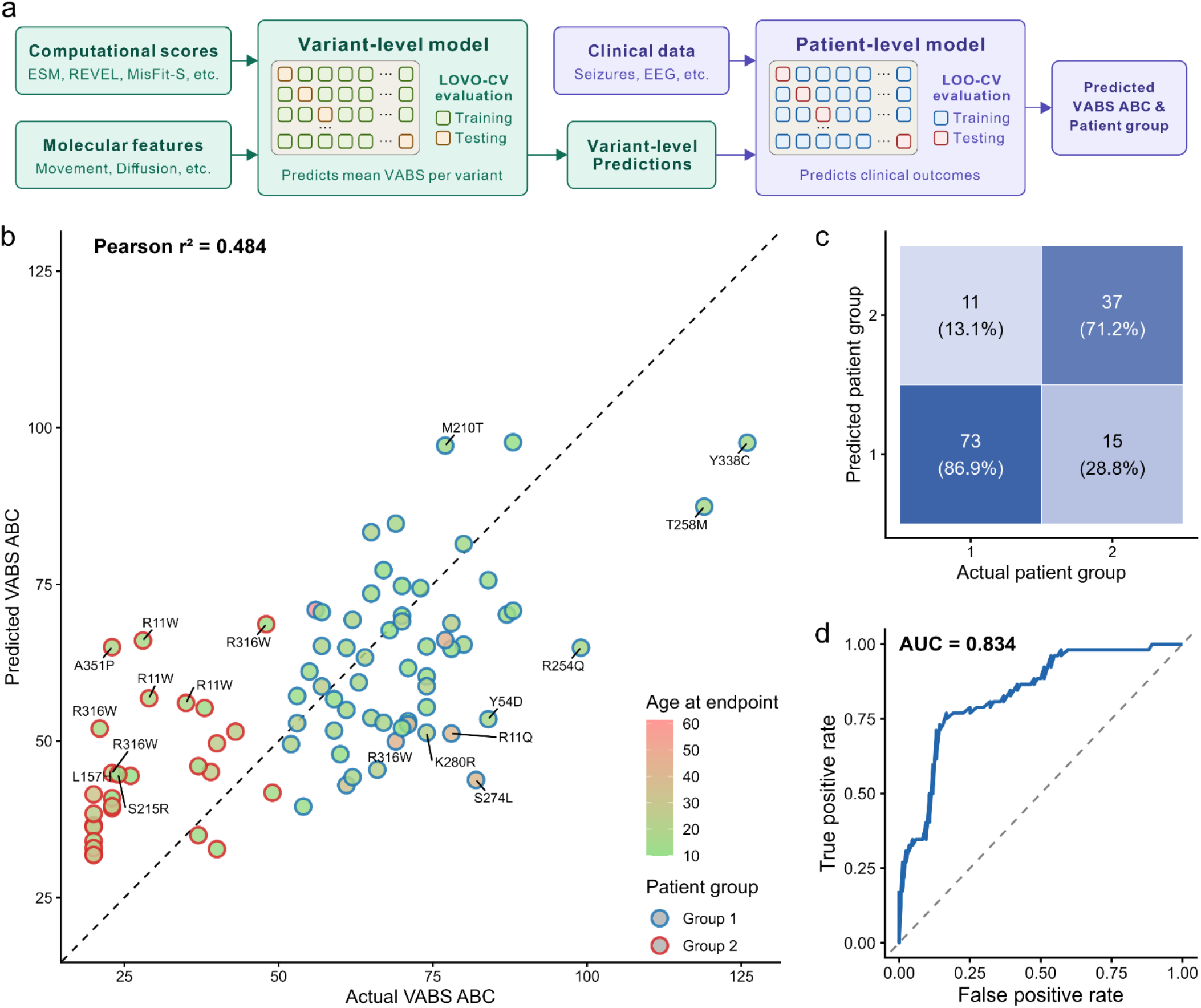
Random Forest model predicts most recent VABS ABC score and patient group. **(a)** Architecture of the two-stage machine learning model. The variant-level model takes computational scores and molecular phenotype data as input to predict mean VABS ABC per variant. The predicted results are then passed to the patient-level model along with clinical phenotype data to predict most recent VABS ABC score and patient group assignment. **(b)** Predicted most recent VABS ABC scores are significantly correlated with actual most recent VABS ABC scores (Pearson r² = 0.484) in patients aged ≥ 10 years at endpoint. Points are colored by age at endpoint and outlined by patient group. Variants are labeled for patients whose predicted and actual VABS ABC scores differ by more than 20. **(c)** Confusion matrix showing model performance in predicting patient group. Note that the number of patients included in group classification is smaller than that for VABS ABC prediction, as patients with unclassified patient group are excluded. (d) Receiver operating characteristic (ROC) curve for patient group prediction, demonstrating moderate-to-good discriminative ability with an AUC of 0.834.

For VABS ABC prediction, the patient-level model achieved a Pearson r = 0.647 between predicted and actual values across all ages (**Supplementary Fig. 8**), a substantial improvement over the variant-level model alone (r = 0.444). Among patients aged ≥10 years at endpoint—the age range over which trajectories stabilize—predicted scores explained 48.4% of the variance in actual scores (r² = 0.484, **Fig. 4b**), demonstrating the model’s ability to capture inter-individual variability in adaptive behavior. For patient group classification, the model correctly classified 86.9% of Group 1 patients (73 of 84) and 71.2% of Group 2 patients (37 of 52) (**Fig. 4c**). The ROC curve yielded an AUC of 0.834, indicating good discriminative ability (**Fig. 4d**). Variants carried by multiple patients (e.g., R316W, R11W, R11Q) showed within-variant variability in prediction accuracy, reflecting genuine inter-individual heterogeneity beyond variant identity. Patient-level predicted and actual VABS ABC scores and group assignments for all individuals are provided in **Supplementary Table 4**.

Ablation analysis confirmed that each feature category contributes independently to model performance (**Supplementary Tables 5 and 6**). At the variant level, removing either molecular features or computational scores reduced predictive accuracy (r = 0.376 and 0.343, respectively, vs. 0.444 for the full model). At the patient level, removing seizures/EEG features produced the largest performance drop (r = 0.557), and further removing molecular features degraded performance (r = 0.410), confirming that the molecular phenotype provides prognostic information beyond clinical variables alone. The same pattern held for group classification: AUC fell to 0.804 without EEG/seizure features and to 0.753 and 0.732 when molecular features or computational scores were additionally removed. Feature importance ranked dwell time as the top variant-level predictor, followed by MisFit-D, velocity, and MisFit-S. At the patient level, the Stage 1 variant-level prediction was the top regressor and seizures were the top classifier (**Supplementary Fig. 9**). Together, these results show that effective prognostication in KAND requires integrating molecular, computational, and clinical information.

## 3. Discussion

There are more than 7000 rare monogenic disorders [27], often affecting children, and ∼70% involving the nervous system [28]. Providing an accurate prognosis for these conditions remains a formidable challenge owing to their rarity and allelic heterogeneity. KIF1A-associated neurological disorder (KAND) is among the more extensively studied rare neurological disorders, yet it encompasses a remarkably broad clinical spectrum, including hereditary spastic paraplegia, neurodevelopmental delay, optic nerve atrophy, peripheral neuropathy, and seizures [8]. Clinical severity varies widely by genetic variant, ranging from mildly affected individuals who survive into at least middle age to severely affected patients who die within the first decade of life. Cohort-level averages, therefore, provide limited guidance for individual patients, underscoring the unmet need for variant-level prediction of clinical outcomes in KAND.

In this study, we developed an integrated, multi-level framework to predict longitudinal adaptive behavior trajectories in KAND by combining single-molecule motor phenotypes, computational pathogenicity scores, and longitudinal clinical data from 343 patients, we demonstrated that accurate prediction of individual outcomes requires integrating multiple data layers. Our two-stage machine learning model achieved a Pearson r² = 0.484 for VABS regression in patients aged ≥10 years and an AUC = 0.834 for patient group classification under leave-one-variant-out cross-validation, providing a quantitative framework directly relevant to clinical decision-making and clinical trial planning.

VABS ABC scores followed two distinct trajectories after age 10, with one group maintaining relatively stable adaptive function while the other declined. Similar bifurcated trajectories have been observed in other developmental epileptic encephalopathies [29, 30]. This divergence suggests that the period before age 10 may represent a critical window for intervention, as patients at risk of severe decline could benefit most from disease-modifying therapies initiated before trajectories diverge [31].

The strong association between EEG/seizure status and VABS outcomes confirms and extends findings from prior work in smaller KAND cohorts [13]. EEG/seizure status emerged as the single most important clinical predictor in the patient-level model. Whether seizures directly cause neuronal injury or are a marker of a more severe underlying molecular phenotype, remains an open question. Regardless of directionality, these findings have immediate clinical implications: all newly diagnosed KAND patients should receive a baseline EEG, and aggressive seizure management may be particularly important for maximizing adaptive function.

At the molecular level, run length and dwell time emerged as the most clinically relevant single-molecule features. Run length showed the strongest positive correlation with VABS ABC among all molecular features (r = 0.37), while dwell time was the most important predictor in the variant-level model. Run length is a direct readout of cargo delivery efficiency in axons [32, 33], and recent high-resolution cryo-EM structures of KIF1A in complex with microtubules have further illuminated how specific pathogenic variants, including P305L, impair microtubule binding and thereby reduce processivity [5]. Variants characterized by prolonged microtubule dwell time, such as T99K and T99M, were associated with the most severe clinical outcomes, including a 5.8-fold higher mortality rate (16.7% vs. 2.9%) compared to the overall cohort. These rigor-like variants are mechanistically distinct from simple loss-of-function mutations [34], suggesting that allele-targeted strategies to reduce mutant protein expression may be particularly appropriate for this subset. Hierarchical clustering of molecular features into two clusters recapitulated the binary patient group structure, driven primarily by whether the variant retained any processive movement. The correlation between MisFit-S and molecular features such as velocity and run length further demonstrates that computational scores capture mechanistic information, providing a rationale for their inclusion as complementary predictors, particularly for variants lacking experimental single-molecule data.

The two-stage framework demonstrated that molecular, computational, and clinical data each contribute distinct and complementary prognostic information that cannot be fully substituted by any single data type. The variant-level model, trained solely on molecular and computational features, captured a meaningful proportion of variance in VABS outcomes, reflecting the fundamental role of KIF1A motor dysfunction in determining disease severity [33, 35, 36]. The patient-level model substantially improved upon this by incorporating clinical features, particularly EEG/seizure status, highlighting that individual clinical course is shaped by factors beyond variant identity alone, a pattern consistent with the variable expressivity documented across rare monogenic disorders [16]. Ablation analysis confirmed that EEG/seizure status made the largest individual contribution to model performance, while single-molecule motility features and computational scores each provided additional independent predictive value; removing any single category reduced overall accuracy. The model performed best for patients with intermediate VABS scores, with relatively lower accuracy at the extremes of the distribution, likely reflecting limited representation of these subgroups in the current dataset. Given that accurate prediction of disease progression has the greatest clinical urgency, expanding the cohort to include additional Group 2 patients will be a priority for future model refinement.

Beyond KAND, the framework presented here applies to other rare monogenic disorders. Three properties make this generalization plausible. First, the biophysical parameters measured in our assay reflect conserved mechanisms of the kinesin motor domain, and pathogenic variants in other kinesin genes, including KIF5A in hereditary spastic paraplegia [37, 38] and amyotrophic lateral sclerosis [39], KIF21A in congenital fibrosis of the extraocular muscles [40, 41], and KIF11 in microcephaly with chorioretinopathy [42, 43], are similarly amenable to single-molecule motility characterization. Prior work has reported variant-specific motility defects in several of these disorders [44–49]. Second, the framework’s reliance on functional measurement rather than population frequency means it remains applicable to ultra-rare or newly identified variants for which conventional approaches fail. Third, the underlying logic extends in principle to any disease protein family for which a scalable functional assay can be developed, including channelopathies measured by patch-clamp electrophysiology, enzymes measured by functional assays, and structural protein variants assessed by biophysical characterization. Systematic cross-disease validation of this paradigm is an important direction for future work.

This study has several limitations. First, the sample size remains modest, and performance estimates for rare variant subgroups may be unstable, a challenge inherent in rare-disease research where cohort sizes are constrained [27]. Second, single-molecule measurements were performed on mutant homodimers rather than heterodimers. We have previously shown that mutant homodimer motility parameters are significantly correlated with VABS ABC scores, whereas those of heterodimers are not [6], likely because severe mutations abolish motility in homodimers while the wild-type subunit partially compensates in heterodimers. Third, patients carrying highly recurrent variants such as R11W and R316W showed dispersed VABS distributions not fully captured by the current model. Fourth, group assignment in our trajectory-based stratification used a fixed VABS ABC threshold of 50, chosen with reference to the cohort distribution. Future work using sensitivity analyses across nearby thresholds and fully data-driven approaches, such as latent class growth analysis or finite mixture modeling, will further validate the bifurcated trajectory framework. Fifth, the model performance is based on cross-validation within the discovery cohort, and prospective validation in an independent KAND cohort represents the principal remaining methodological gap before clinical deployment.

In summary, VABS trajectories in KAND patients bifurcate into two distinct patterns strongly associated with clinical phenotypes, particularly EEG/seizure status. When a novel *KIF1A* variant is identified, its single-molecule phenotype can be characterized using the smTIRF pipeline described here, and these parameters can be integrated with computational variant-effect scores into our predictive model to generate individualized prognostic estimates. The model is dynamic and can be continuously updated and refined as additional clinical data accumulate. Beyond prognosis, this framework has direct implications for therapeutic development and clinical trial design as disease-modifying treatments advance. Broader adoption of standardized outcomes, particularly the VABS and Growth Scales, in future natural history studies and clinical registries will help expand the variant-outcome dataset and improve model generalizability.

## 4. Methods

### 4.1 Ethics and Participant Recruitment

This study was approved by the Columbia University and Boston Children’s Hospital Institutional Review Boards, and informed consent was obtained from all individuals or their guardians. The study included individuals with *KIF1A* pathogenic or likely pathogenic variants, as defined by the American College of Medical Genetics (ACMG) classification guidelines. All individuals had previously undergone exome sequencing, genome sequencing, or panel gene testing. Clinical genetic test reports were reviewed by a geneticist, and individuals with dual genetic diagnoses affecting the nervous system were excluded.

### 4.2 Clinical Data Collection

Medical data were collected through a telephone interview with a physician or genetic counselor. Follow-up clinical data were collected via online questionnaires completed by caregivers at intervals of at least 12 months and were validated against medical records. Adaptive function was assessed using the Vineland Adaptive Behavior Scales, 3rd Edition (VABS-3), comprehensive parent/caregiver form, administered via the online Pearson Q-global platform. Longitudinal VABS follow-ups were administered at least 12 months apart.

To be included in the machine learning analysis, patients were required to have at least one VABS ABC assessment and either single-molecule phenotype data or computational pathogenicity scores for their variant. Patients without any VABS record were excluded. For deceased patients, age at death and a VABS ABC floor value of 20 (the minimum scale score) were assigned to their most recent endpoint for modeling. Patients who died before age 10 were directly assigned to Group 2, reflecting the severe disease course associated with early mortality. For patients with longitudinal VABS data, the last available assessment served as the endpoint for all analyses.

### 4.3 Generation of Plasmids for KIF1A Constructs

A plasmid encoding a truncated KIF1A construct (KIF1A [*Homo sapiens*, aa 1–393]–leucine zipper–SNAPf–EGFP–6His) was used as the template for all constructs in this study. KIF1A mutations were introduced using Q5 mutagenesis (New England Biolabs, #E0554S) and confirmed by Sanger sequencing.

### 4.4 Protein Expression in *E. coli*

Each plasmid was transformed into BL21-CodonPlus(DE3)-RIPL competent cells (Agilent Technologies, #230280). A single colony was inoculated into 1 mL of terrific broth (TB) containing 50 µg/mL carbenicillin and 50 µg/mL chloramphenicol and incubated overnight at 37 °C with shaking. The culture was then transferred to 400 mL of TB containing 2 µg/mL of each antibiotic, grown at 37 °C for 5 h, cooled to 16 °C for 1 h, and induced with 0.1 mM IPTG at 16 °C overnight. Cells were harvested by centrifugation at 3,000 × g for 10 min at 4 °C. The pellet was resuspended in B-PER™ Complete Bacterial Protein Extraction Reagent (ThermoFisher Scientific, #89821) supplemented with 2 mM MgCl₂, 1 mM EGTA, 1 mM DTT, 0.1 mM ATP, and 2 mM PMSF, then flash-frozen in liquid nitrogen.

### 4.5 Protein Purification

The frozen cell pellet was thawed at 37 °C and nutated at room temperature for 20 min to lyse the cells. The lysate was clarified by centrifugation at 80,000 rpm (260,000 × g) for 10 min, then passed through 500 µL of Roche cOmplete His-Tag purification resin (Millipore Sigma, #5893682001). The resin was washed with wash buffer (50 mM HEPES, 300 mM KCl, 2 mM MgCl₂, 1 mM EGTA, 1 mM DTT, 1 mM PMSF, 0.1 mM ATP, 0.1% Pluronic F-127 [w/v], 10% glycerol, pH 7.2) and incubated with 10 µM SNAP-Cell TMR-Star (New England Biolabs, #S9105S) at room temperature for 10 min. After additional washing, protein was eluted with elution buffer containing 150 mM imidazole, flash-frozen, and stored at −80 °C.

### 4.6 Microtubule-Binding and Release (MTBR) Assay

To remove inactive motors before single-molecule TIRF assays, an MTBR assay was performed. Fifty µL of eluted protein was buffer-exchanged into low-salt buffer (30 mM HEPES, 50 mM KCl, 2 mM MgCl₂, 1 mM EGTA, 1 mM DTT, 1 mM AMP-PNP, 10 µM taxol, 0.1% Pluronic F-127 [w/v], 10% glycerol) using a 0.5 mL Zeba spin desalting column (ThermoFisher Scientific, #89882). After warming to room temperature, 5 µL of 5 mg/mL paclitaxel-stabilized microtubules was added, and the mixture was incubated for 2 min. The mixture was centrifuged at 45,000 rpm (80,000 × g) for 5 min at room temperature. The pellet was washed twice with 20 µL low-salt buffer and resuspended in 50 µL high-salt release buffer (30 mM HEPES, 300 mM KCl, 2 mM MgCl₂, 1 mM EGTA, 1 mM DTT, 10 µM paclitaxel, 3 mM ATP, 0.1% Pluronic F-127 [w/v], 10% glycerol). Microtubules were removed by centrifugation at 40,000 rpm (60,000 × g) for 5 min. The supernatant containing active motors was aliquoted, flash-frozen, and stored at −80 °C.

### 4.7 Single-Molecule TIRF Motility Assay

MTBR fractions were used for single-molecule TIRF assays with motor dilutions adjusted to achieve appropriate surface densities. A flow chamber was assembled from a glass slide (Fisher #12-550-123), an ethanol-cleaned coverslip (Zeiss #474030-9000-000), and two parafilm strips. All incubations were performed at room temperature. The chamber was sequentially incubated with 0.5 mg/mL BSA-biotin (10 min), blocked with blocking buffer (80 mM PIPES, 2 mM MgCl₂, 1 mM EGTA, 10 µM paclitaxel, 1% Pluronic F-127 [w/v], pH 6.8, 10 min), incubated with 0.25 mg/mL streptavidin (10 min), and loaded with 0.02 mg/mL Cy5-and biotin-labeled microtubules (1 min). The MTBR motor fraction was diluted in motility buffer (80 mM PIPES, 2 mM MgCl₂, 1 mM EGTA, 1 mM DTT, 10 µM paclitaxel, 0.5% Pluronic F-127 [w/v], 2 mM ATP, 5 mg/mL BSA, 1 mg/mL α-casein, gloxy oxygen scavenging system, 10% glycerol, pH 7.2) and introduced into the chamber. Images were acquired at 200 ms per frame for 600 frames using an appropriate TIRF microscope setup, and kymographs were analyzed using a custom MATLAB software (R2025a).

### 4.8 Computational Metrics of KIF1A variants

Multiple computational scores were obtained for all KIF1A variants. REVEL (rare exome variant ensemble learner) is an ensemble score that integrates multiple functional prediction tools, with higher scores indicating greater likelihood of pathogenicity (range 0–1) [24]. gMVP (graphical missense variant pathogenicity predictor) predicts the functional impact of missense variants using a gradient boosting model trained on ClinVar and gnomAD data, with higher scores indicating a greater predicted pathogenicity (range 0–1) [25]. ESM (evolutionary scale model)-1b scores represent the log-likelihood ratio of the mutant versus wild-type amino acid derived from a protein language model trained on 250 million protein sequences; more negative values indicate greater deviation from evolutionary expectation and thus more damaging functional consequences [17]. AlphaMissense predicts the pathogenicity of missense variants using a deep learning model based on the AlphaFold2 architecture, with scores ranging from 0 to 1, where higher values indicate greater predicted pathogenicity [26]. MisFit-D and MisFit-S are two complementary scores from the MisFit framework [18]. MisFit-D estimates the molecular-level damage caused by a missense variant, with higher values indicating greater disruption of protein function. MisFit-S estimates the heterozygous selection coefficient, reflecting the fitness cost of carrying the variant in human populations, with higher values indicating stronger purifying selection and greater predicted pathogenicity.

### 4.9 Trajectory-Based Patient Stratification

To classify patients into distinct functional trajectory groups, we developed a prediction-based stratification approach using longitudinal VABS ABC scores. For each patient, two features were extracted: the first recorded VABS ABC score (*VABS*_1*st*_) and the rate of change (β) estimated via linear regression across all available observations. The predicted VABS ABC score at age 10 was calculated as:

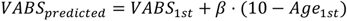

For patients with at least one evaluation at age ≥ 10 years, the minimum recorded VABS ABC score after age 10 was used directly. For patients with evaluations only before age 10 and at least two time points, the predicted value was used. Patients with a single observation were excluded from classification.

Group assignment followed these rules: patients whose predicted or observed VABS ABC at age 10 fell below a predefined threshold (50, based on the VABS ABC distribution across all patients) were assigned to Group 2; all remaining patients were assigned to Group 1. Additionally, patients who died before age 10 were assigned directly to Group 2, and patients whose all recorded VABS ABC scores exceeded 100 (at least twice) were assigned directly to Group 1. Patients with only a single pre-age-10 observation were designated as unclassified.

### 4.10 Hierarchical Clustering of Molecular Phenotype Data

All missense variants’ single-molecule motility features, including movement status (binary), diffusion status (binary), velocity, run length, dwell time, and conservation score, were min-max scaled to [0, 1] before clustering. Hierarchical clustering was performed using Ward’s D2 method with Euclidean distances. The optimal number of clusters (k = 2) was determined by visual inspection of the dendrogram and the biological interpretability of the resulting clusters. The clustering and visualization were implemented in R using the ComplexHeatmap package (version 2.24.1).

### 4.11 Statistical Association Analyses

Univariate linear regression was used to assess associations between each predictor variable and the most recent VABS ABC score or its change, with adjustment for sex and age at the most recent VABS evaluation, where applicable. Categorical variables (clinical phenotypes) were encoded as binary indicators. Pairwise Pearson correlations among molecular features, computational scores, and clinical outcomes were computed and visualized as heatmaps using the ComplexHeatmap package. Between-group comparisons of continuous variables were performed using the Wilcoxon rank-sum test, and comparisons of categorical variables were made using Fisher’s exact test. All statistical analyses were performed in R version 4.5.0. A two-sided P value < 0.05 was considered statistically significant.

### 4.12 Two-Stage Machine Learning Prediction Framework

We developed a two-stage machine learning model in R using the caret package (version 7.0.1), with Random Forest (RF) as the base algorithm for both stages, to predict VABS ABC scores and patient group membership. For patients with longitudinal VABS assessments, the most recent available VABS ABC score was used as the primary outcome measure in all analyses.

Stage 1 (Variant-Level Model): The first stage predicts the variant-level mean VABS ABC from molecular phenotype features (biophysical motor parameters) and computational pathogenicity scores. For recurrent variants, patient records were aggregated into a single row per unique variant by computing the mean of the last available VABS ABC scores (for the regression task) or the mean of numeric patient group assignments (for the classification task) across all carriers. The first-stage model was trained and evaluated using Leave-One-Variant-Out Cross-Validation (LOVO-CV), in which each unique variant is held out as the test set in turn while the model is trained on all remaining variants. This design ensures that performance reflects true out-of-sample generalization across variants rather than across individual patients. Cross-validated variant-level predictions were then propagated back to all patients sharing the corresponding variant.

Stage 2 (Patient-Level Model): The second stage predicts patient-level outcomes from a combined feature set that includes Stage 1 variant-level predictions and patient-level clinical predictors, such as age at VABS assessment, EEG/seizure status, and other clinical variables. For the VABS ABC regression task, Stage 2 produces a continuous predicted score; for the patient group classification task, it produces a binary group assignment. Stage 2 was evaluated using Leave-One-Out Cross-Validation (LOO-CV) at the patient level, restricted to patients with a valid Stage 1 prediction.

For both stages, the model hyperparameter “mtry” was selected via repeated 5-fold cross-validation on the full training set before the LOVO-CV and LOO-CV evaluation loops, and the optimal configuration was applied uniformly across all held-out folds. Regression performance was assessed using Root Mean Square Error (RMSE), Mean Absolute Error (MAE), coefficient of determination (R²), and Pearson correlation coefficient, while classification performance was assessed using accuracy, balanced accuracy, sensitivity, specificity, and area under the ROC curve (AUC), computed using the pROC package (v1.19.0.1). To assess the independent contributions of different feature categories, we performed ablation analysis by systematically removing molecular features, computational pathogenicity scores, age, and EEG/seizure features individually and in combination from their respective model stages, and computed performance differences relative to the full model (delta metrics) for each configuration. Feature importance was assessed using the percentage increase in mean squared error for regression tasks and the mean decrease in accuracy for classification tasks, both derived from the full-data model after cross-validation.

## Data Availability

The data sets used and/or analyzed during the current study are available from the corresponding author on request.

## Code Availability

All software used in this study is publicly available. The code for major figures and analysis can be found at https://github.com/liwenxing2016/KIF1A_prediction.

## Author Contribution

W.L. curated clinical data, annotated KIF1A variants with computational scores, developed the machine learning framework, performed all data analyses, and interpreted the results. J.W., C.C, C.T., C.A.L. and W.K.C. collected clinical data. L.R. performed single-molecule experiments and analyzed the data. Y.S., W.K.C., and A.G. conceived and supervised the project. W.L., L.R., Y.S., W.K.C., and A.G. wrote the manuscript. Y.S., W.K.C., and A.G. secured funding. All authors reviewed and approved the final manuscript.

## Supplementary Tables and Figures

**Supplementary Fig. 1.**
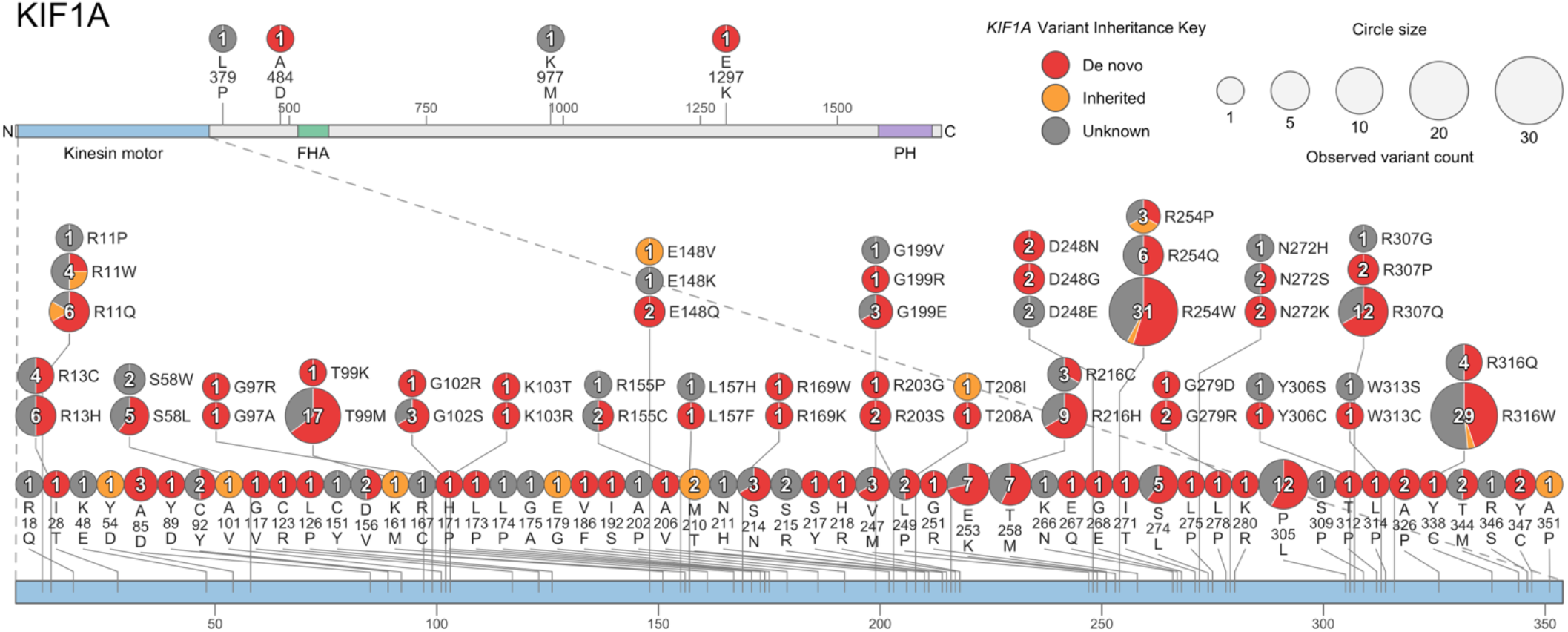
KIF1A missense variants in the cohort. Each circle represents a unique missense variant in KIF1A (UniProt Q12756; NM_001244008.2), sized by the number of unrelated individuals carrying it, and drawn as a pie chart color-coded by inheritance. Inheritance was grouped as de novo, inherited (maternally or paternally inherited), or unknown (inheritance unknown, unreported, or likely gonadal mosaicism). Positions with more than one distinct change are shown as vertically stacked circles. The top shows the full-length protein with its Kinesin motor, FHA, and PH domains; the bottom is an enlargement of the Kinesin motor domain. Only missense variants are shown: 291 individuals with 110 unique variants; 22 individuals with non-missense or unmappable variants are not included.

**Supplementary Fig. 2.**
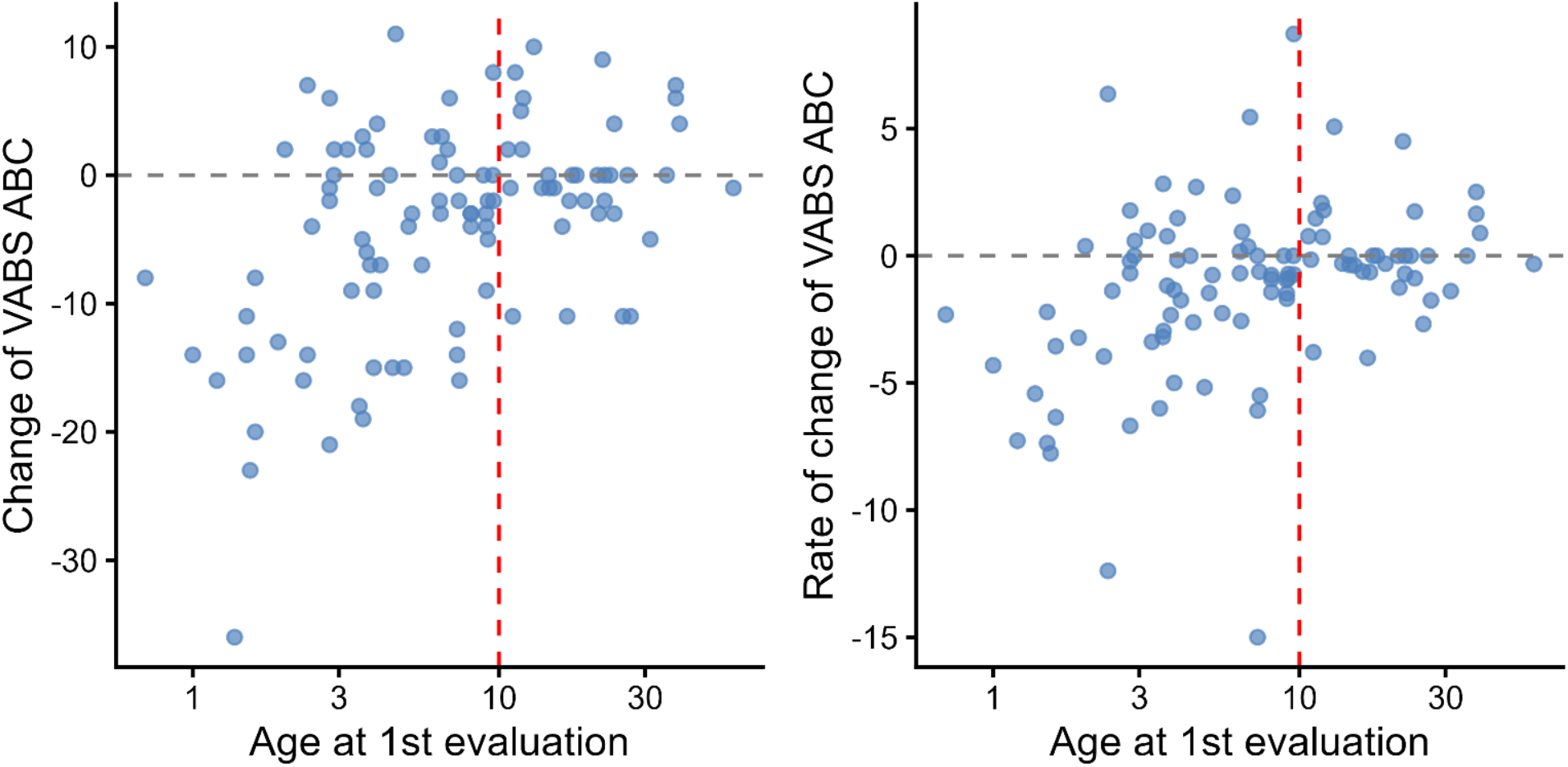
Change in VABS ABC score and rate of change relative to age at first evaluation. Each point represents one patient with longitudinal VABS assessments. The left panel shows the absolute change in VABS ABC score between first and last assessment, and the right panel shows the rate of change per year. The red dashed line indicates age 10. Both the magnitude and rate of VABS ABC change are more variable in patients with a first evaluation before age 10, and become more stable thereafter, supporting the use of the last available VABS ABC score as the primary outcome measure and age 10 as a meaningful inflection point for trajectory analysis.

**Supplementary Fig. 3.**
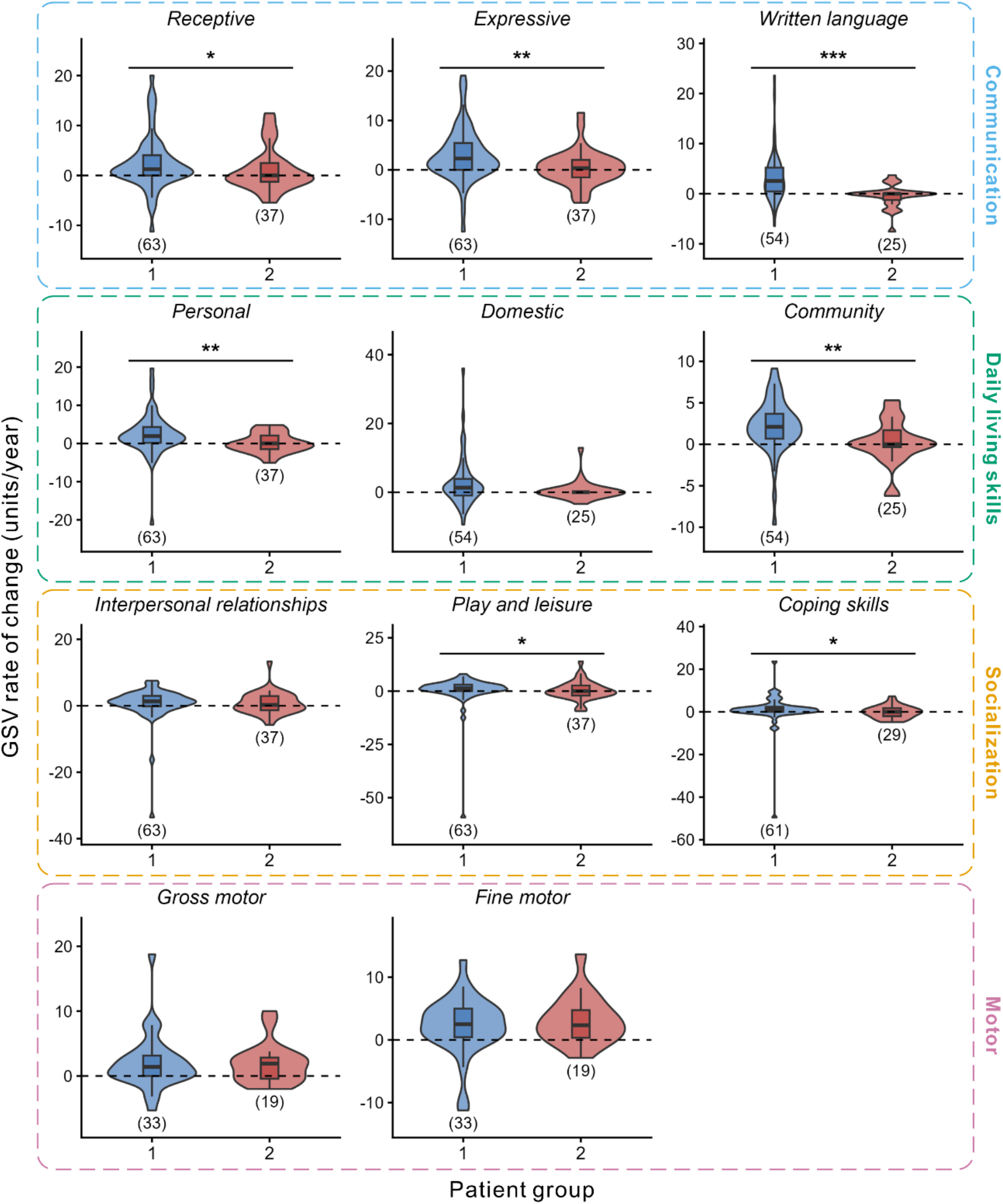
Growth scale value (GSV) rate of change across subdomains by patient group. Violin plots show the distribution of GSV rate of change (ΔGSV/year, calculated as the difference between most recent and first available GSV divided by the follow-up interval in years) for Group 1 and Group 2 patients (described in Fig. 1) across all 11 Vineland-3 subdomains. Dashed horizontal line indicates zero rate of change. Numbers in parentheses indicate sample size. Between-group comparisons by Wilcoxon rank-sum test. \**P* < 0.05, \*\**P* < 0.01, \*\*\**P* < 0.001.

**Supplementary Fig. 4.**
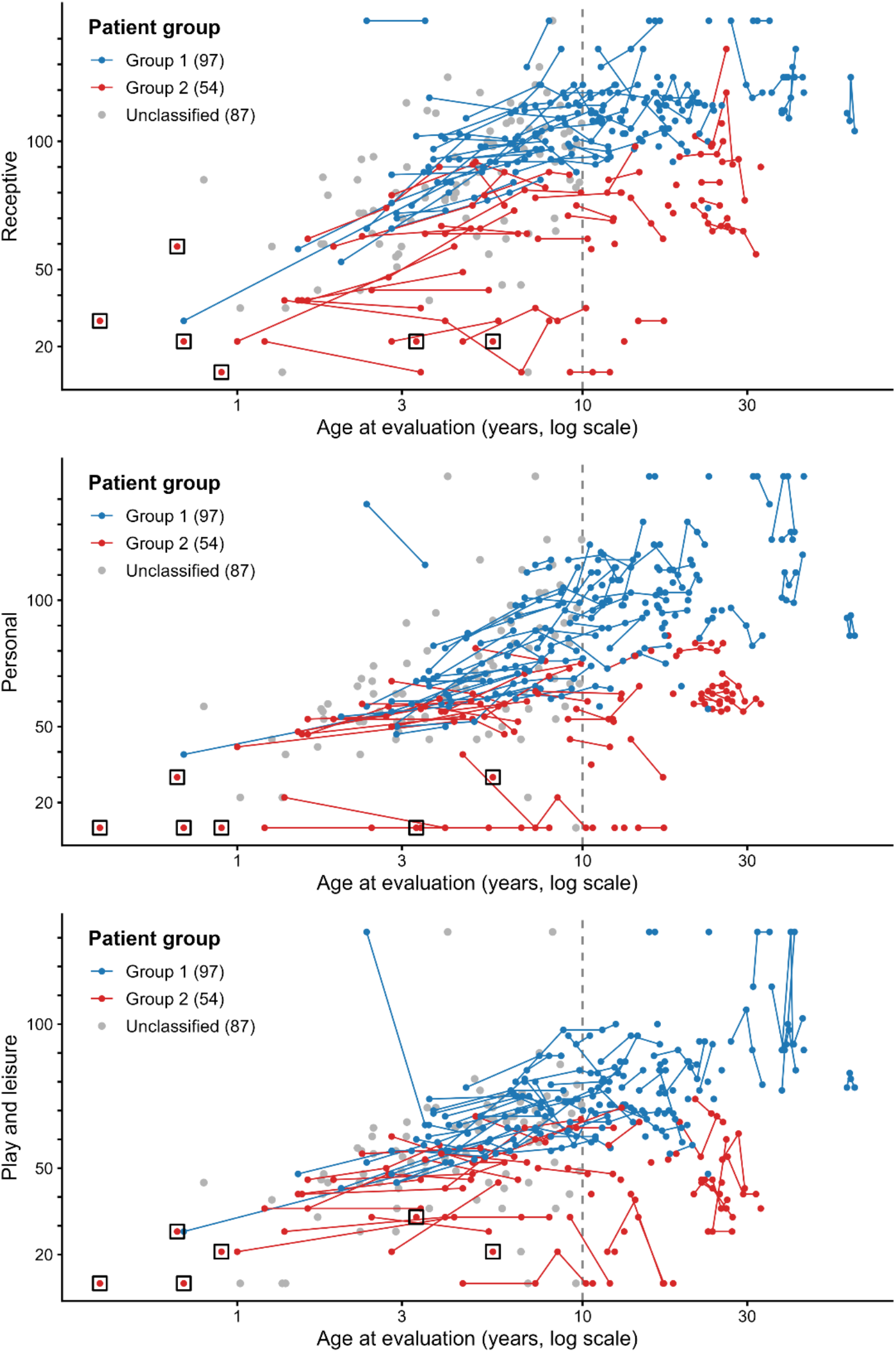
Longitudinal growth scale value (GSV) trajectories across representative Vineland-3 subdomains. Trajectories are shown for three representative subdomains: Receptive (Communication), Personal (Daily Living Skills), and Play and leisure (Socialization). Each line connects longitudinal observations from a single person, unconnected points represent patients with a single assessment. Patient groups were defined in Fig. 1. Hollow squares indicate patients who died before age 10. In all three subdomains, Group 1 patients showed continued GSV accumulation, indicating ongoing acquisition of new skills, whereas Group 2 patients showed minimal GSV change, indicating a stall in skill acquisition, with trajectories remaining at lower absolute values across the observation period.

**Supplementary Fig. 5.**
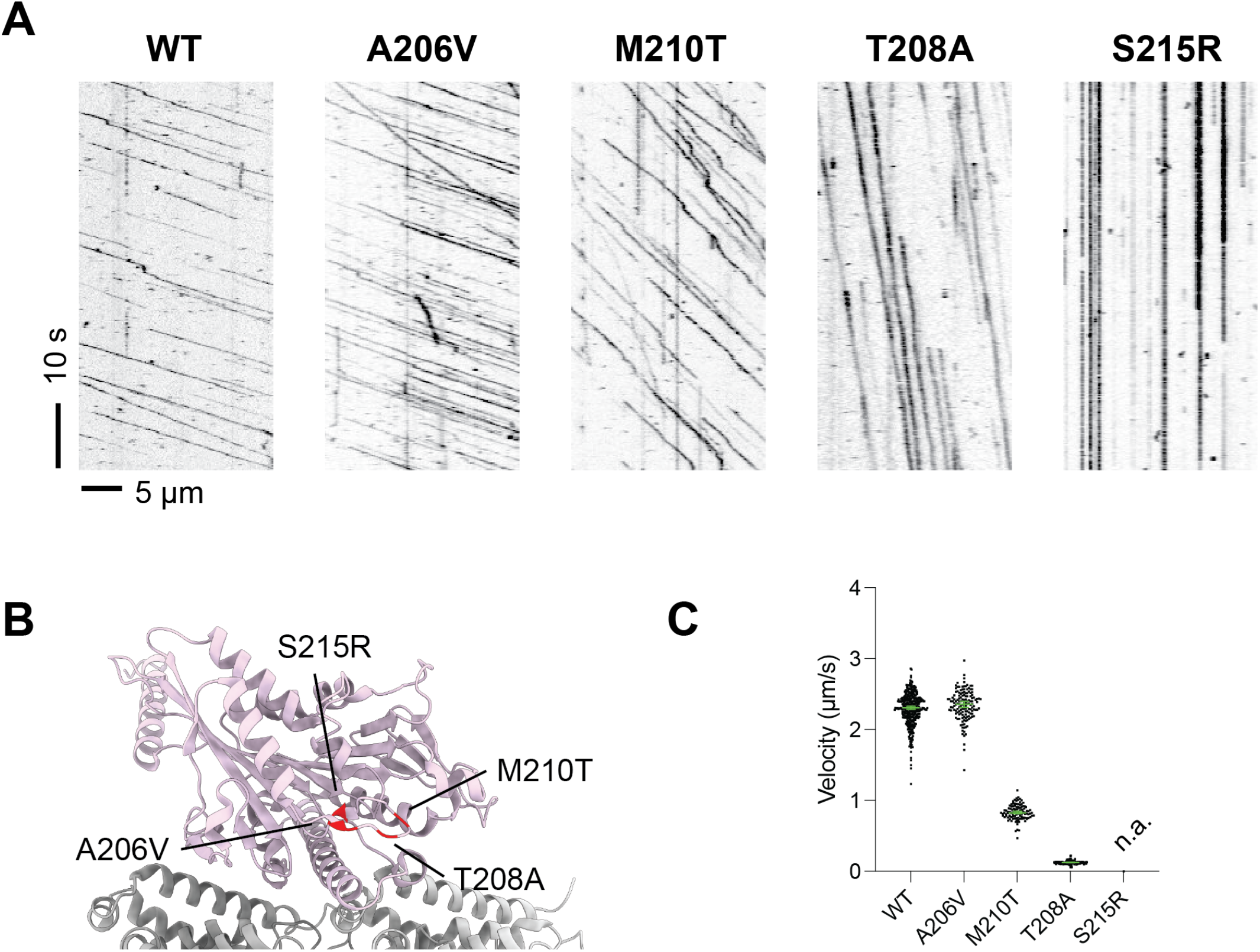
Representative single-molecule motility phenotypes of KIF1A variants. **(a)** Kymographs from smTIRF microscopy showing the motility of wild-type (WT) KIF1A and four disease-associated variants at the switch-I loop. Diagonal lines indicate processive movement; vertical lines indicate stationary microtubule binding; absence of lines indicates no microtubule interaction. Time scale bar: 10 s; spatial scale bar: 5 µm. (b) Structural mapping of the four variants onto the KIF1A motor domain, highlighting their spatial proximity within the switch-I loop (red). (c) Velocity distributions for each variant. S215R showed no processive movement and velocity was not applicable (n.a.). Despite mapping to adjacent residues, the four variants exhibit markedly divergent motility phenotypes, ranging from near-wild-type motility (A206V) to reduced velocity (M210T), stationary microtubule binding (T208A), and complete loss of motility (S215R), illustrating that structural proximity does not predict functional consequence and underscoring the necessity of single-molecule characterization for each variant individually.

**Supplementary Fig. 6.**
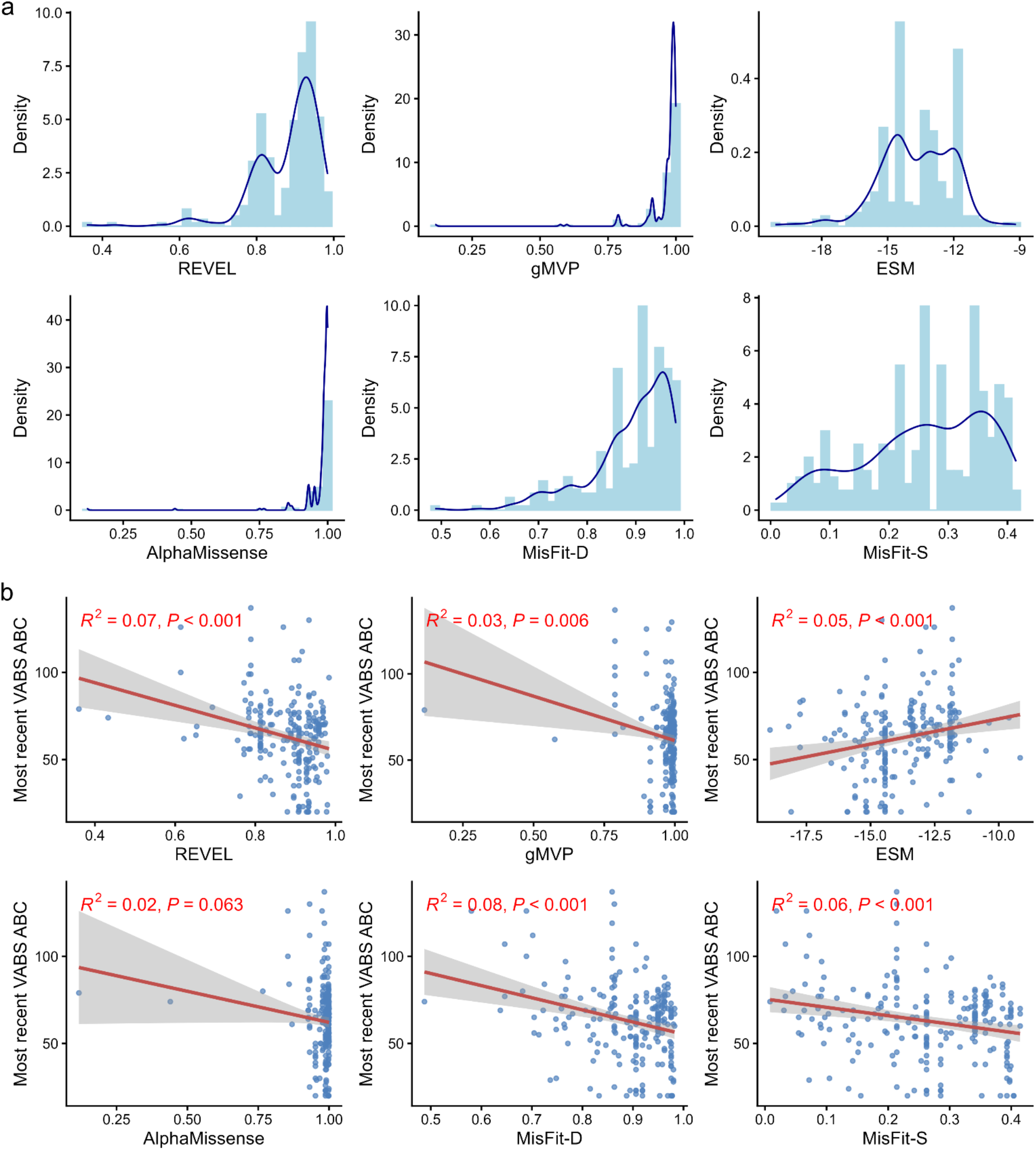
Distribution and clinical associations of computational scores across all KIF1A variants. (A) Density plots showing the distribution of six computational scores across all patients included in this study, reflecting the score distribution weighted by variant recurrence. (B) Scatter plots showing the correlation between each computational score and most recent VABS ABC score in all patients with available computational scores. Red lines indicate linear regression fits with 95% confidence intervals. *R*² and *P* values shown are from Pearson correlation analysis.

**Supplementary Fig. 7.**
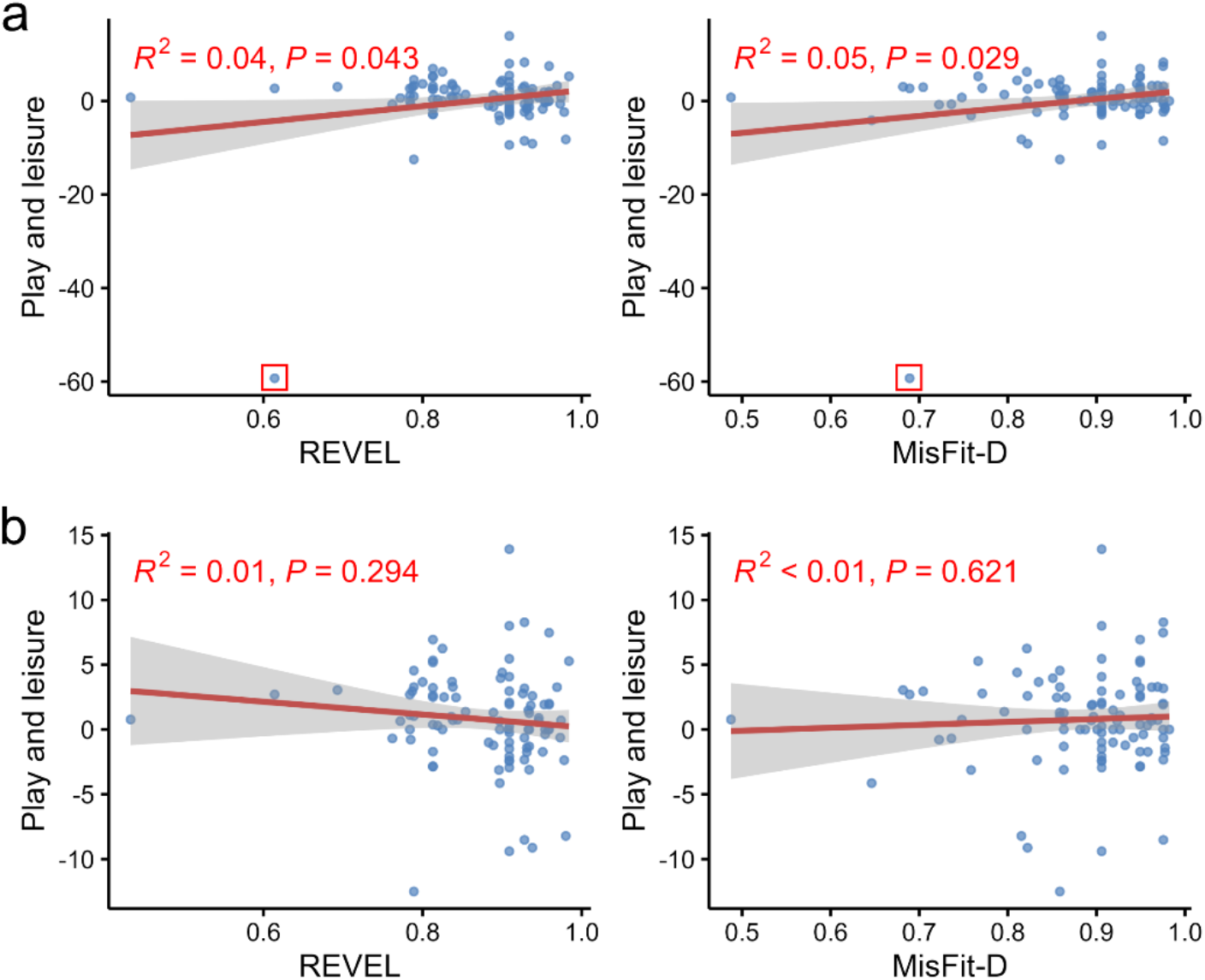
Sensitivity analysis for the association between computational pathogenicity scores and GSV rate of change in the play and leisure subdomain. Scatter plots show the associations between REVEL (left) and MisFit-D (right) with GSV rate of change in the play and leisure subdomain in the full cohort (a) and after removal of a single high-influence observation (b, red square in a). The positive associations observed in the full cohort (REVEL: R² = 0.04, P = 0.043; MisFit-D: R² = 0.05, P = 0.029) were not significant after removal of this observation (REVEL: R² = 0.01, P = 0.294; MisFit-D: R² < 0.01, P = 0.621), indicating that the positive correlations between computational pathogenicity scores and GSV rate of change in the play and leisure subdomain observed in Fig. 3c are driven by a single influential observation and do not reflect a true biological relationship.

**Supplementary Fig. 8.**
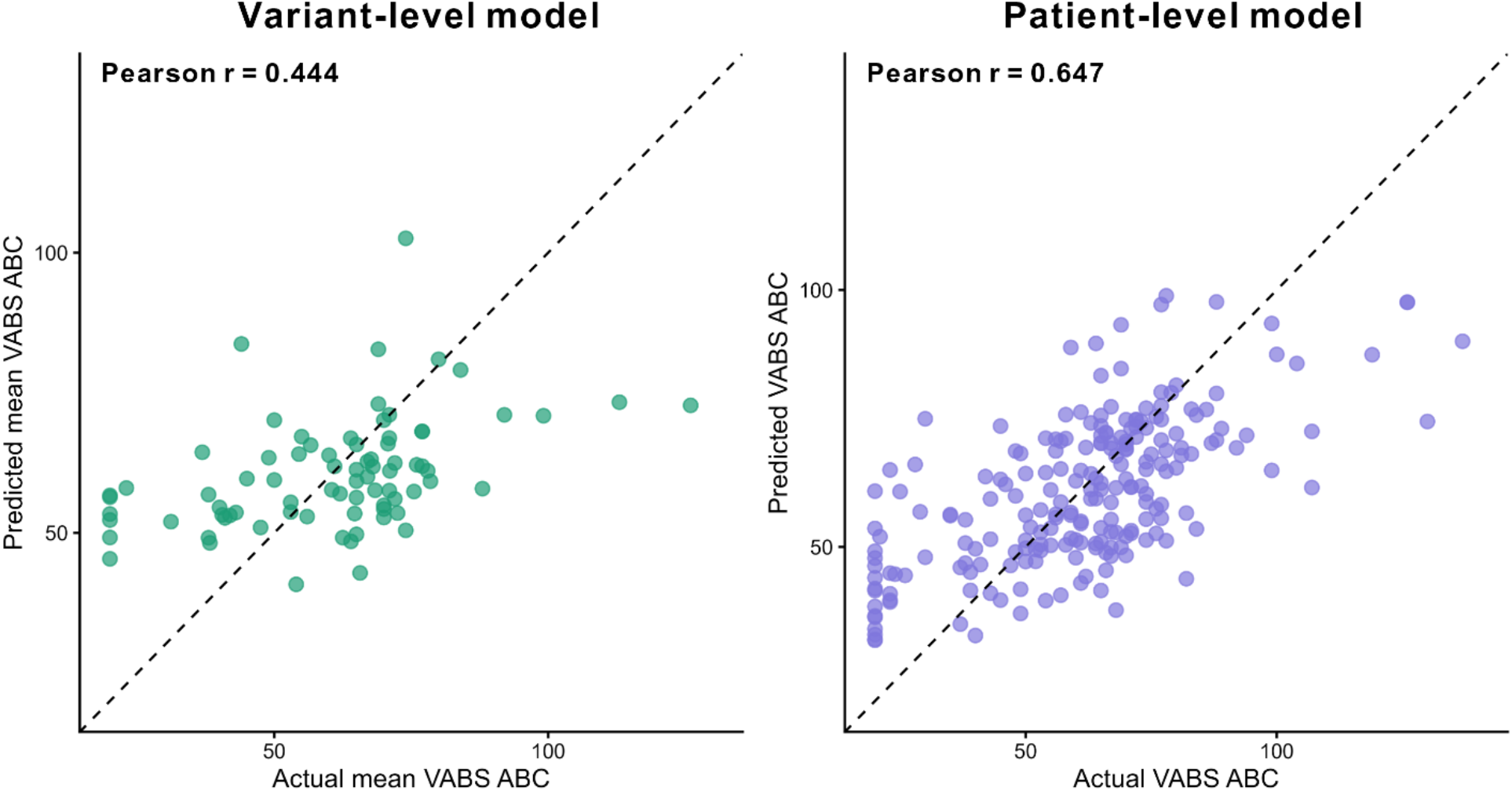
Predicted versus actual VABS ABC scores for the variant-level and patient-level models. Scatter plots show the correlation between predicted and actual VABS ABC scores for the variant-level model (left, green) and patient-level model (right, purple). Each point represents one variant (left) or one patient (right).

**Supplementary Fig. 9.**
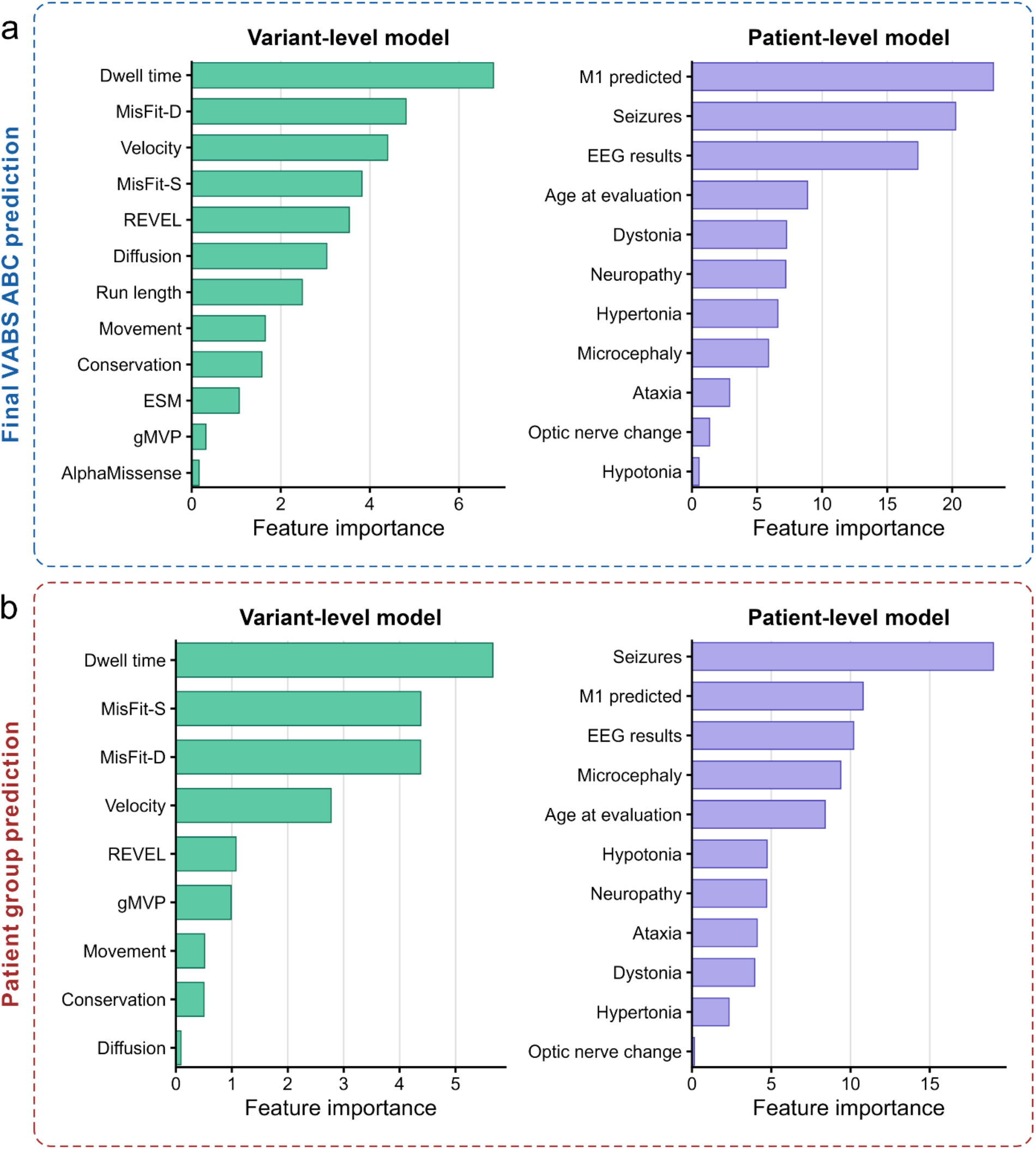
Feature importance in variant-level and patient-level models for most recent VABS ABC prediction (a) and patient group classification (b). Feature importance for the variant-level model (green) is measured as the percentage increase in mean squared error upon permutation of each feature, and for the patient-level model (purple) as the mean decrease in accuracy. Only features with positive importance values are shown. In both tasks, dwell time was the most important feature in the variant-level model, followed by MisFit-D, velocity, and MisFit-S. In the patient-level model, the Stage 1 variant-level predictions (M1 predicted) ranked highest for VABS ABC regression, while seizures ranked highest for patient group classification.

**Supplementary Table 1.**
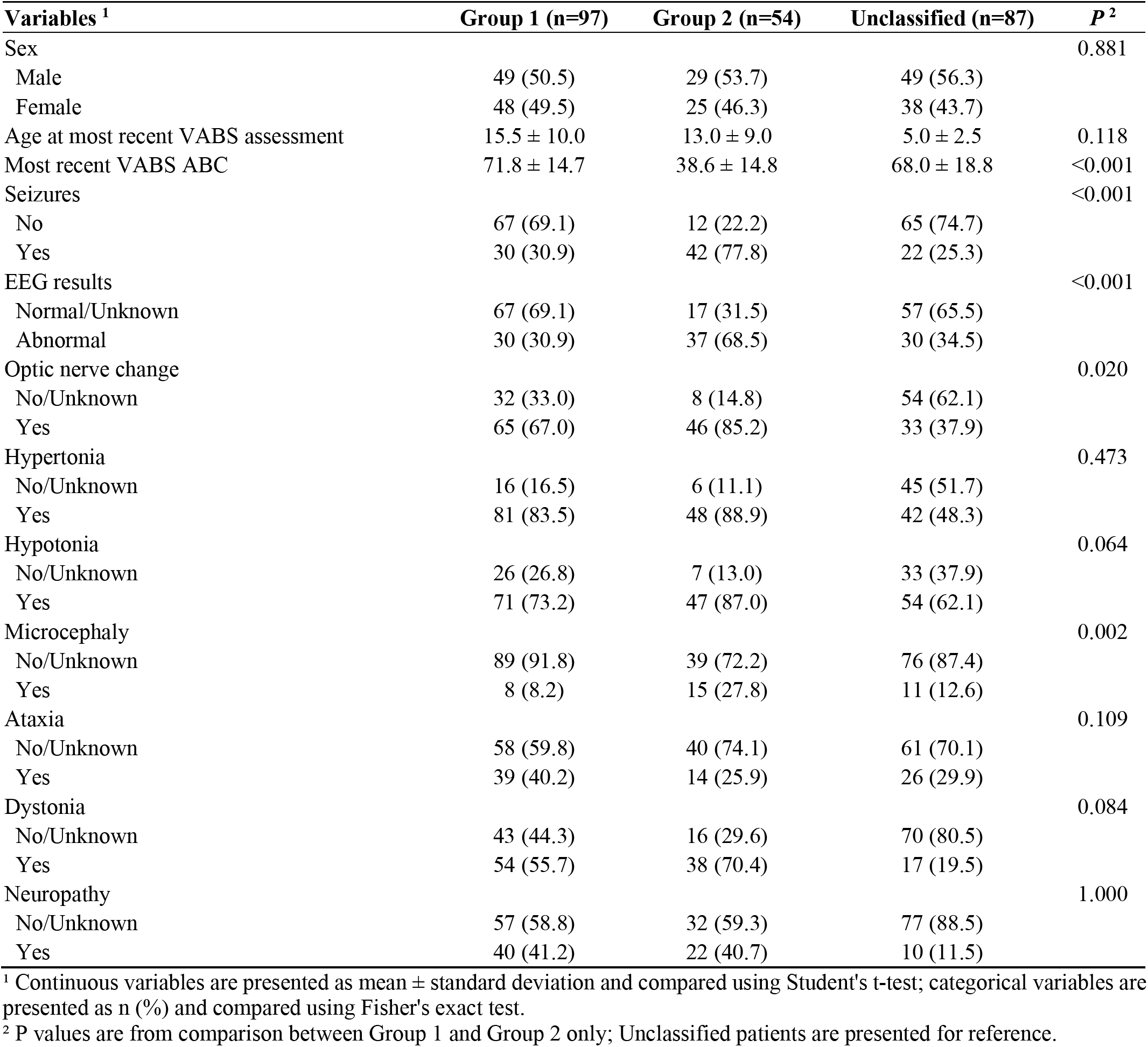
Baseline clinical features of Group 1, Group 2, and Unclassified patients.

| Variables <sup>1</sup> | Group 1 (n=97) | Group 2 (n=54) | Unclassified (n=87) | P <sup>2</sup> |
| --- | --- | --- | --- | --- |
| Sex |  |  |  | 0.881 |
| Male | 49 (50.5) | 29 (53.7) | 49 (56.3) |  |
| Female | 48 (49.5) | 25 (46.3) | 38 (43.7) |  |
| Age at most recent VABS assessment | 15.5 ± 10.0 | 13.0 ± 9.0 | 5.0 ± 2.5 | 0.118 |
| Most recent VABS ABC | 71.8 ± 14.7 | 38.6 ± 14.8 | 68.0 ± 18.8 | <0.001 |
| Seizures |  |  |  | <0.001 |
| No | 67 (69.1) | 12 (22.2) | 65 (74.7) |  |
| Yes | 30 (30.9) | 42 (77.8) | 22 (25.3) |  |
| EEG results |  |  |  | <0.001 |
| Normal/Unknown | 67 (69.1) | 17 (31.5) | 57 (65.5) |  |
| Abnormal | 30 (30.9) | 37 (68.5) | 30 (34.5) |  |
| Optic nerve change |  |  |  | 0.020 |
| No/Unknown | 32 (33.0) | 8 (14.8) | 54 (62.1) |  |
| Yes | 65 (67.0) | 46 (85.2) | 33 (37.9) |  |
| Hypertonia |  |  |  | 0.473 |
| No/Unknown | 16 (16.5) | 6 (11.1) | 45 (51.7) |  |
| Yes | 81 (83.5) | 48 (88.9) | 42 (48.3) |  |
| Hypotonia |  |  |  | 0.064 |
| No/Unknown | 26 (26.8) | 7 (13.0) | 33 (37.9) |  |
| Yes | 71 (73.2) | 47 (87.0) | 54 (62.1) |  |
| Microcephaly |  |  |  | 0.002 |
| No/Unknown | 89 (91.8) | 39 (72.2) | 76 (87.4) |  |
| Yes | 8 (8.2) | 15 (27.8) | 11 (12.6) |  |
| Ataxia |  |  |  | 0.109 |
| No/Unknown | 58 (59.8) | 40 (74.1) | 61 (70.1) |  |
| Yes | 39 (40.2) | 14 (25.9) | 26 (29.9) |  |
| Dystonia |  |  |  | 0.084 |
| No/Unknown | 43 (44.3) | 16 (29.6) | 70 (80.5) |  |
| Yes | 54 (55.7) | 38 (70.4) | 17 (19.5) |  |
| Neuropathy |  |  |  | 1.000 |
| No/Unknown | 57 (58.8) | 32 (59.3) | 77 (88.5) |  |
| Yes | 40 (41.2) | 22 (40.7) | 10 (11.5) |  |

<sup>1</sup> Continuous variables are presented as mean ± standard deviation and compared using Student's t-test; categorical variables are presented as n (%) and compared using Fisher's exact test.
<sup>2</sup> P values are from comparison between Group 1 and Group 2 only; Unclassified patients are presented for reference.

**Supplementary Table 2.** Univariate linear regression of the association between clinical phenotypes, computational scores, and molecular features with most recent VABS ABC in all patients.

| Variables | N | Mean (SD) | Crude |  | Adjusted<sup>1</sup> |  |
| --- | --- | --- | --- | --- | --- | --- |
|  |  |  | Beta (SE) | P | Beta (SE) | P |
| *Clinical phenotypes (lifetime)* |  |  |  |  |  |  |
| Seizures |  |  |  |  |  |  |
| No | 144 | 70.22 ± 19.34 | Reference |  | Reference |  |
| Yes | 94 | 51.66 ± 18.45 | -18.56 (2.52) | <0.001 | -18.00 (2.52) | <0.001 |
| EEG results |  |  |  |  |  |  |
| Normal/Unknown | 141 | 70.05 ± 20.13 | Reference |  | Reference |  |
| Abnormal | 97 | 52.48 ± 17.76 | -17.57 (2.53) | <0.001 | -17.63 (2.50) | <0.001 |
| Optic nerve change |  |  |  |  |  |  |
| No/Unknown | 94 | 71.21 ± 21.01 | Reference |  | Reference |  |
| Yes | 144 | 57.46 ± 19.23 | -13.75 (2.65) | <0.001 | -12.90 (2.68) | <0.001 |
| Hypertonia |  |  |  |  |  |  |
| No/Unknown | 67 | 64.75 ± 16.06 | Reference |  | Reference |  |
| Yes | 171 | 62.16 ± 22.67 | -2.58 (3.03) | 0.395 | -0.45 (3.13) | 0.886 |
| Hypotonia |  |  |  |  |  |  |
| No/Unknown | 66 | 70.94 ± 23.92 | Reference |  | Reference |  |
| Yes | 172 | 59.80 ± 18.98 | -11.14 (2.96) | <0.001 | -11.27 (2.93) | <0.001 |
| Microcephaly |  |  |  |  |  |  |
| No/Unknown | 204 | 64.43 ± 21.25 | Reference |  | Reference |  |
| Yes | 34 | 53.68 ± 17.13 | -10.75 (3.84) | 0.006 | -11.90 (3.80) | 0.002 |
| Ataxia |  |  |  |  |  |  |
| No/Unknown | 159 | 62.98 ± 22.21 | Reference |  | Reference |  |
| Yes | 79 | 62.71 ± 18.52 | -0.27 (2.90) | 0.925 | 0.53 (2.88) | 0.855 |
| Dystonia |  |  |  |  |  |  |
| No/Unknown | 129 | 67.24 ± 21.19 | Reference |  | Reference |  |
| Yes | 109 | 57.74 ± 19.71 | -9.50 (2.67) | <0.001 | -8.26 (2.76) | 0.003 |
| Neuropathy |  |  |  |  |  |  |
| No/Unknown | 166 | 65.28 ± 20.87 | Reference |  | Reference |  |
| Yes | 72 | 57.38 ± 20.45 | -7.91 (2.93) | 0.007 | -5.76 (3.25) | 0.077 |
| *Computational scores* |  |  |  |  |  |  |
| REVEL | 226 |  | -64.84 (16.06) | <0.001 | -63.56 (15.92) | <0.001 |
| gMVP | 226 |  | -51.48 (18.54) | 0.006 | -50.21 (18.38) | 0.007 |
| ESM | 223 |  | 2.90 (0.85) | 0.001 | 2.76 (0.85) | 0.001 |
| AlphaMissense | 226 |  | -35.66 (19.08) | 0.063 | -34.30 (18.91) | 0.071 |
| MisFit-D | 223 |  | -69.62 (16.18) | <0.001 | -69.21 (16.01) | <0.001 |
| MisFit-S | 223 |  | -47.90 (13.17) | <0.001 | -46.91 (13.05) | <0.001 |
| *Molecular features* |  |  |  |  |  |  |
| Movement |  |  |  |  |  |  |
| No | 76 | 55.11 ± 19.18 | Reference |  | Reference |  |
| Yes | 137 | 66.36 ± 21.20 | 11.25 (2.93) | <0.001 | 10.93 (2.89) | <0.001 |
| Diffusion |  |  |  |  |  |  |
| No | 127 | 64.54 ± 21.69 | Reference |  | Reference |  |
| Yes | 86 | 59.10 ± 20.02 | -5.43 (2.94) | 0.066 | -5.90 (2.89) | 0.042 |
| Velocity | 213 |  | 9.17 (2.11) | <0.001 | 9.14 (2.08) | <0.001 |
| Run length | 213 |  | 3.72 (0.64) | <0.001 | 3.71 (0.63) | <0.001 |
| Dwell time (log) | 213 |  | -2.97 (1.32) | 0.026 | -3.32 (1.30) | 0.011 |
| Conservation | 213 |  | -33.31 (7.68) | <0.001 | -32.79 (7.55) | <0.001 |

<sup>1</sup> P values were adjusted for sex at birth and age at most recent VABS evaluation. SD: standard deviation, SE: standard error.

**Supplementary Table 3.**
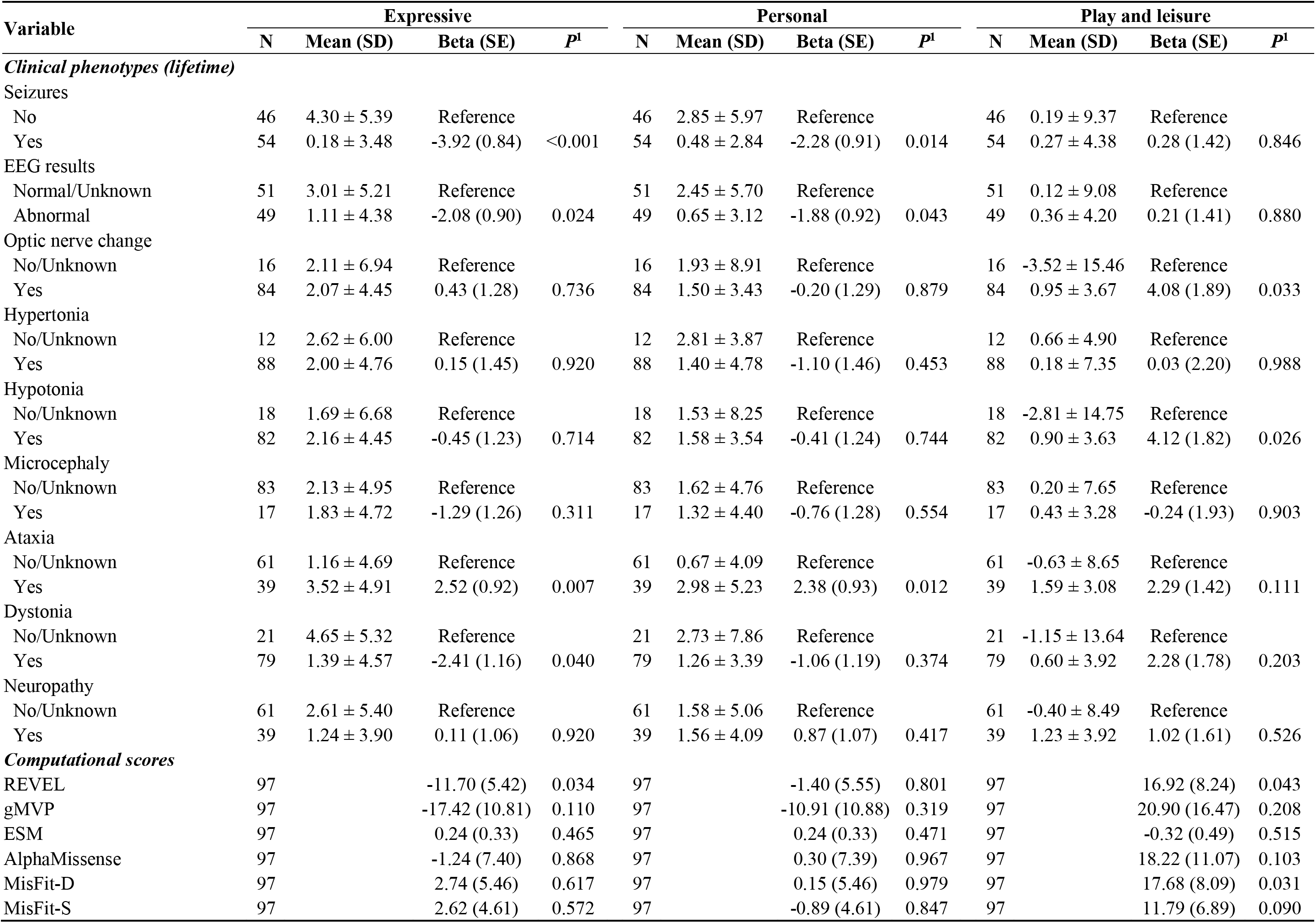
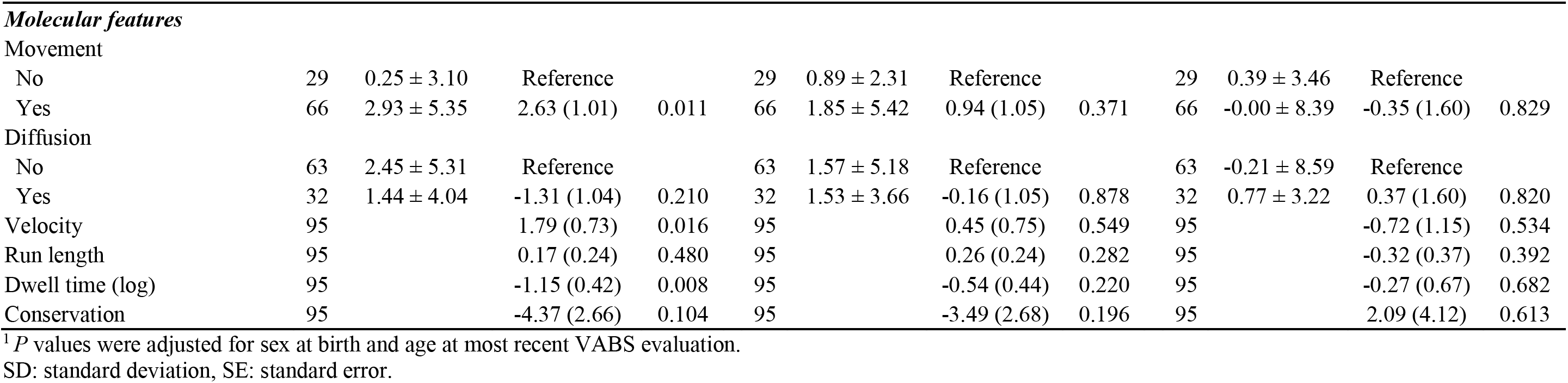
Univariate linear regression of the association between clinical phenotypes, pathogenicity scores, and molecular features with growth scale value rate of change across representative Vineland-3 subdomains.

| Variable | Expressive |  |  |  | Personal |  |  |  | Play and leisure |  |  |  |
| --- | --- | --- | --- | --- | --- | --- | --- | --- | --- | --- | --- | --- |
|  | N | Mean (SD) | Beta (SE) | P<sup>†</sup> | N | Mean (SD) | Beta (SE) | P<sup>†</sup> | N | Mean (SD) | Beta (SE) | P<sup>†</sup> |
| ***Clinical phenotypes (lifetime)*** |  |  |  |  |  |  |  |  |  |  |  |  |
| Seizures |  |  |  |  |  |  |  |  |  |  |  |  |
| No | 46 | 4.30 ± 5.39 | Reference |  | 46 | 2.85 ± 5.97 | Reference |  | 46 | 0.19 ± 9.37 | Reference |  |
| Yes | 54 | 0.18 ± 3.48 | -3.92 (0.84) | <0.001 | 54 | 0.48 ± 2.84 | -2.28 (0.91) | 0.014 | 54 | 0.27 ± 4.38 | 0.28 (1.42) | 0.846 |
| EEG results |  |  |  |  |  |  |  |  |  |  |  |  |
| Normal/Unknown | 51 | 3.01 ± 5.21 | Reference |  | 51 | 2.45 ± 5.70 | Reference |  | 51 | 0.12 ± 9.08 | Reference |  |
| Abnormal | 49 | 1.11 ± 4.38 | -2.08 (0.90) | 0.024 | 49 | 0.65 ± 3.12 | -1.88 (0.92) | 0.043 | 49 | 0.36 ± 4.20 | 0.21 (1.41) | 0.880 |
| Optic nerve change |  |  |  |  |  |  |  |  |  |  |  |  |
| No/Unknown | 16 | 2.11 ± 6.94 | Reference |  | 16 | 1.93 ± 8.91 | Reference |  | 16 | -3.52 ± 15.46 | Reference |  |
| Yes | 84 | 2.07 ± 4.45 | 0.43 (1.28) | 0.736 | 84 | 1.50 ± 3.43 | -0.20 (1.29) | 0.879 | 84 | 0.95 ± 3.67 | 4.08 (1.89) | 0.033 |
| Hypertonia |  |  |  |  |  |  |  |  |  |  |  |  |
| No/Unknown | 12 | 2.62 ± 6.00 | Reference |  | 12 | 2.81 ± 3.87 | Reference |  | 12 | 0.66 ± 4.90 | Reference |  |
| Yes | 88 | 2.00 ± 4.76 | 0.15 (1.45) | 0.920 | 88 | 1.40 ± 4.78 | -1.10 (1.46) | 0.453 | 88 | 0.18 ± 7.35 | 0.03 (2.20) | 0.988 |
| Hypotonia |  |  |  |  |  |  |  |  |  |  |  |  |
| No/Unknown | 18 | 1.69 ± 6.68 | Reference |  | 18 | 1.53 ± 8.25 | Reference |  | 18 | -2.81 ± 14.75 | Reference |  |
| Yes | 82 | 2.16 ± 4.45 | -0.45 (1.23) | 0.714 | 82 | 1.58 ± 3.54 | -0.41 (1.24) | 0.744 | 82 | 0.90 ± 3.63 | 4.12 (1.82) | 0.026 |
| Microcephaly |  |  |  |  |  |  |  |  |  |  |  |  |
| No/Unknown | 83 | 2.13 ± 4.95 | Reference |  | 83 | 1.62 ± 4.76 | Reference |  | 83 | 0.20 ± 7.65 | Reference |  |
| Yes | 17 | 1.83 ± 4.72 | -1.29 (1.26) | 0.311 | 17 | 1.32 ± 4.40 | -0.76 (1.28) | 0.554 | 17 | 0.43 ± 3.28 | -0.24 (1.93) | 0.903 |
| Ataxia |  |  |  |  |  |  |  |  |  |  |  |  |
| No/Unknown | 61 | 1.16 ± 4.69 | Reference |  | 61 | 0.67 ± 4.09 | Reference |  | 61 | -0.63 ± 8.65 | Reference |  |
| Yes | 39 | 3.52 ± 4.91 | 2.52 (0.92) | 0.007 | 39 | 2.98 ± 5.23 | 2.38 (0.93) | 0.012 | 39 | 1.59 ± 3.08 | 2.29 (1.42) | 0.111 |
| Dystonia |  |  |  |  |  |  |  |  |  |  |  |  |
| No/Unknown | 21 | 4.65 ± 5.32 | Reference |  | 21 | 2.73 ± 7.86 | Reference |  | 21 | -1.15 ± 13.64 | Reference |  |
| Yes | 79 | 1.39 ± 4.57 | -2.41 (1.16) | 0.040 | 79 | 1.26 ± 3.39 | -1.06 (1.19) | 0.374 | 79 | 0.60 ± 3.92 | 2.28 (1.78) | 0.203 |
| Neuropathy |  |  |  |  |  |  |  |  |  |  |  |  |
| No/Unknown | 61 | 2.61 ± 5.40 | Reference |  | 61 | 1.58 ± 5.06 | Reference |  | 61 | -0.40 ± 8.49 | Reference |  |
| Yes | 39 | 1.24 ± 3.90 | 0.11 (1.06) | 0.920 | 39 | 1.56 ± 4.09 | 0.87 (1.07) | 0.417 | 39 | 1.23 ± 3.92 | 1.02 (1.61) | 0.526 |
| ***Computational scores*** |  |  |  |  |  |  |  |  |  |  |  |  |
| REVEL | 97 |  | -11.70 (5.42) | 0.034 | 97 |  | -1.40 (5.55) | 0.801 | 97 |  | 16.92 (8.24) | 0.043 |
| gMVP | 97 |  | -17.42 (10.81) | 0.110 | 97 |  | -10.91 (10.88) | 0.319 | 97 |  | 20.90 (16.47) | 0.208 |
| ESM | 97 |  | 0.24 (0.33) | 0.465 | 97 |  | 0.24 (0.33) | 0.471 | 97 |  | -0.32 (0.49) | 0.515 |
| AlphaMissense | 97 |  | -1.24 (7.40) | 0.868 | 97 |  | 0.30 (7.39) | 0.967 | 97 |  | 18.22 (11.07) | 0.103 |
| MisFit-D | 97 |  | 2.74 (5.46) | 0.617 | 97 |  | 0.15 (5.46) | 0.979 | 97 |  | 17.68 (8.09) | 0.031 |
| MisFit-S | 97 |  | 2.62 (4.61) | 0.572 | 97 |  | -0.89 (4.61) | 0.847 | 97 |  | 11.79 (6.89) | 0.090 |
| Movement |  |  |  |  |  |  |  |  |  |  |  |  |
| No | 29 | 0.25 ± 3.10 | Reference |  | 29 | 0.89 ± 2.31 | Reference |  | 29 | 0.39 ± 3.46 | Reference |  |
| Yes | 66 | 2.93 ± 5.35 | 2.63 (1.01) | 0.011 | 66 | 1.85 ± 5.42 | 0.94 (1.05) | 0.371 | 66 | -0.00 ± 8.39 | -0.35 (1.60) | 0.829 |
| Diffusion |  |  |  |  |  |  |  |  |  |  |  |  |
| No | 63 | 2.45 ± 5.31 | Reference |  | 63 | 1.57 ± 5.18 | Reference |  | 63 | -0.21 ± 8.59 | Reference |  |
| Yes | 32 | 1.44 ± 4.04 | -1.31 (1.04) | 0.210 | 32 | 1.53 ± 3.66 | -0.16 (1.05) | 0.878 | 32 | 0.77 ± 3.22 | 0.37 (1.60) | 0.820 |
| Velocity | 95 |  | 1.79 (0.73) | 0.016 | 95 |  | 0.45 (0.75) | 0.549 | 95 |  | -0.72 (1.15) | 0.534 |
| Run length | 95 |  | 0.17 (0.24) | 0.480 | 95 |  | 0.26 (0.24) | 0.282 | 95 |  | -0.32 (0.37) | 0.392 |
| Dwell time (log) | 95 |  | -1.15 (0.42) | 0.008 | 95 |  | -0.54 (0.44) | 0.220 | 95 |  | -0.27 (0.67) | 0.682 |
| Conservation | 95 |  | -4.37 (2.66) | 0.104 | 95 |  | -3.49 (2.68) | 0.196 | 95 |  | 2.09 (4.12) | 0.613 |

<sup>1</sup> *P* values were adjusted for sex at birth and age at most recent VABS evaluation.
SD: standard deviation, SE: standard error.

**Supplementary Table 4.**
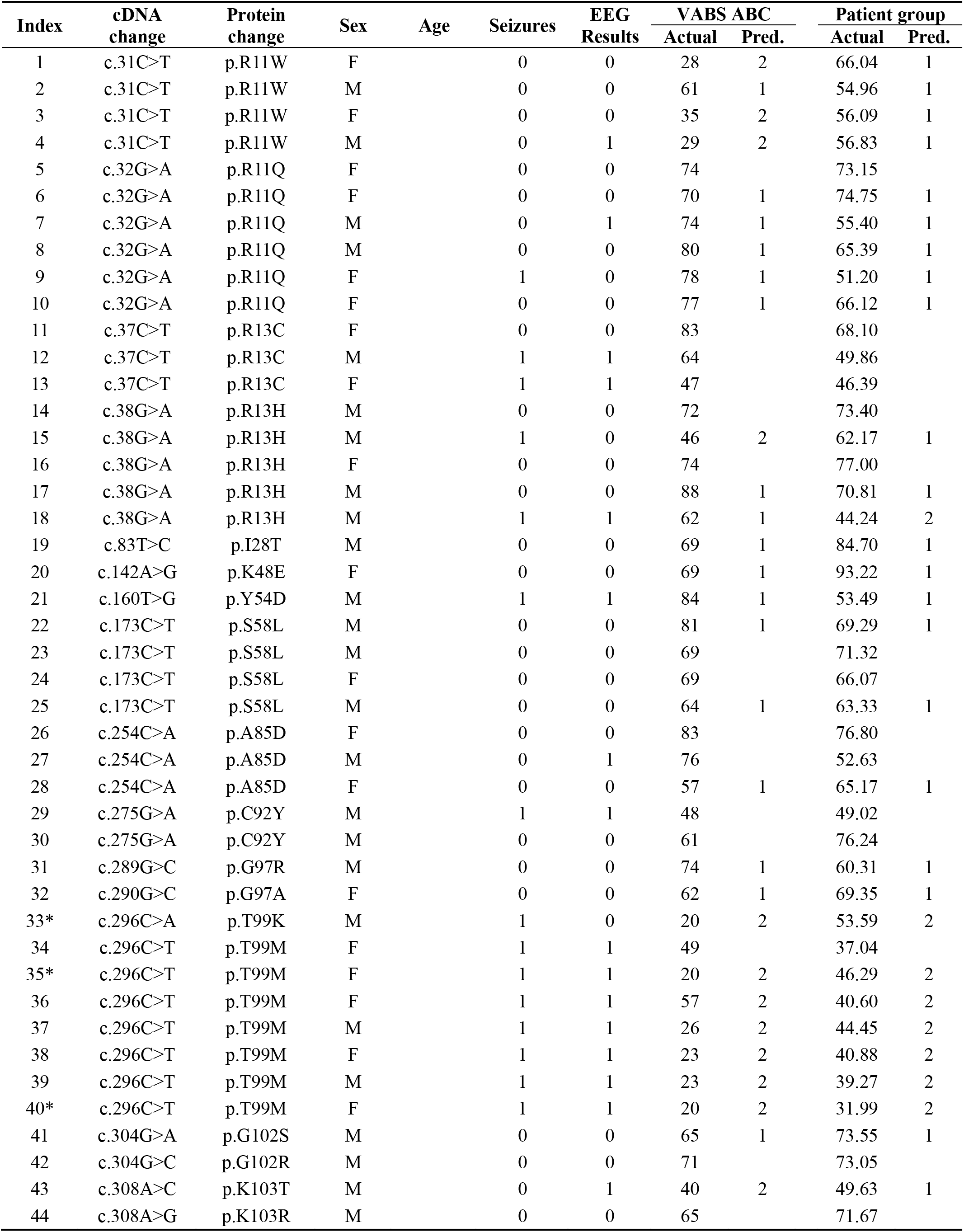
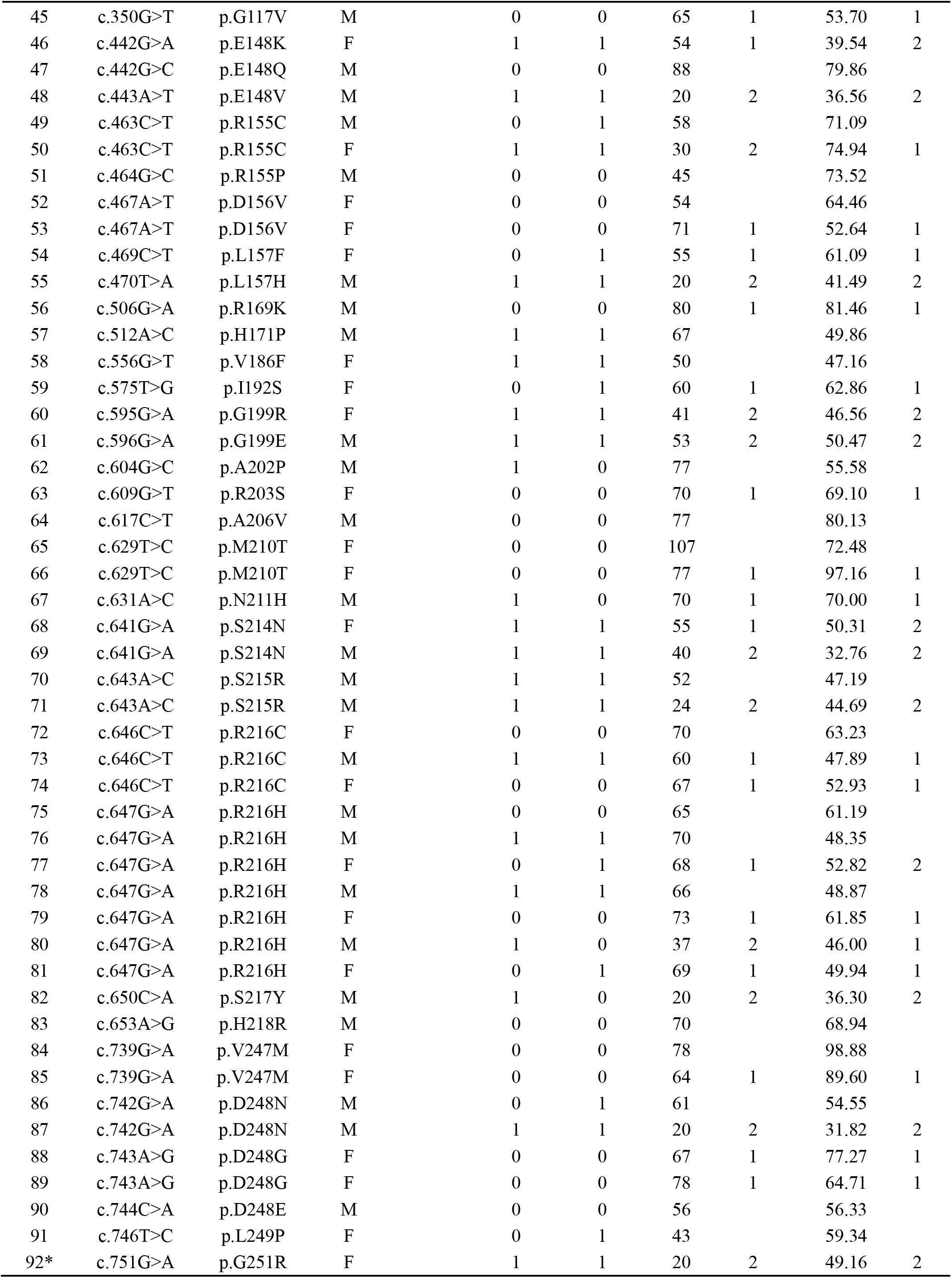
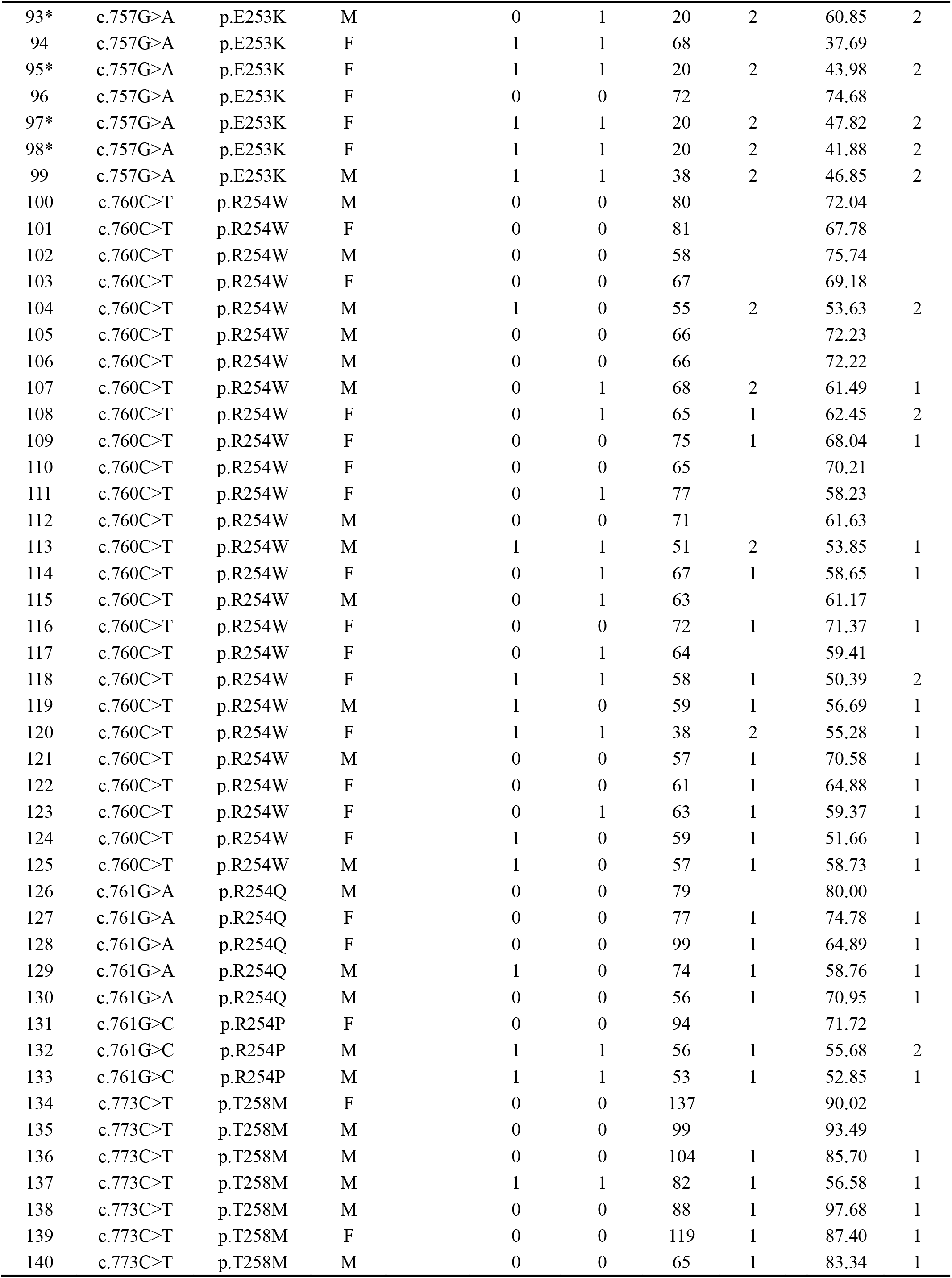
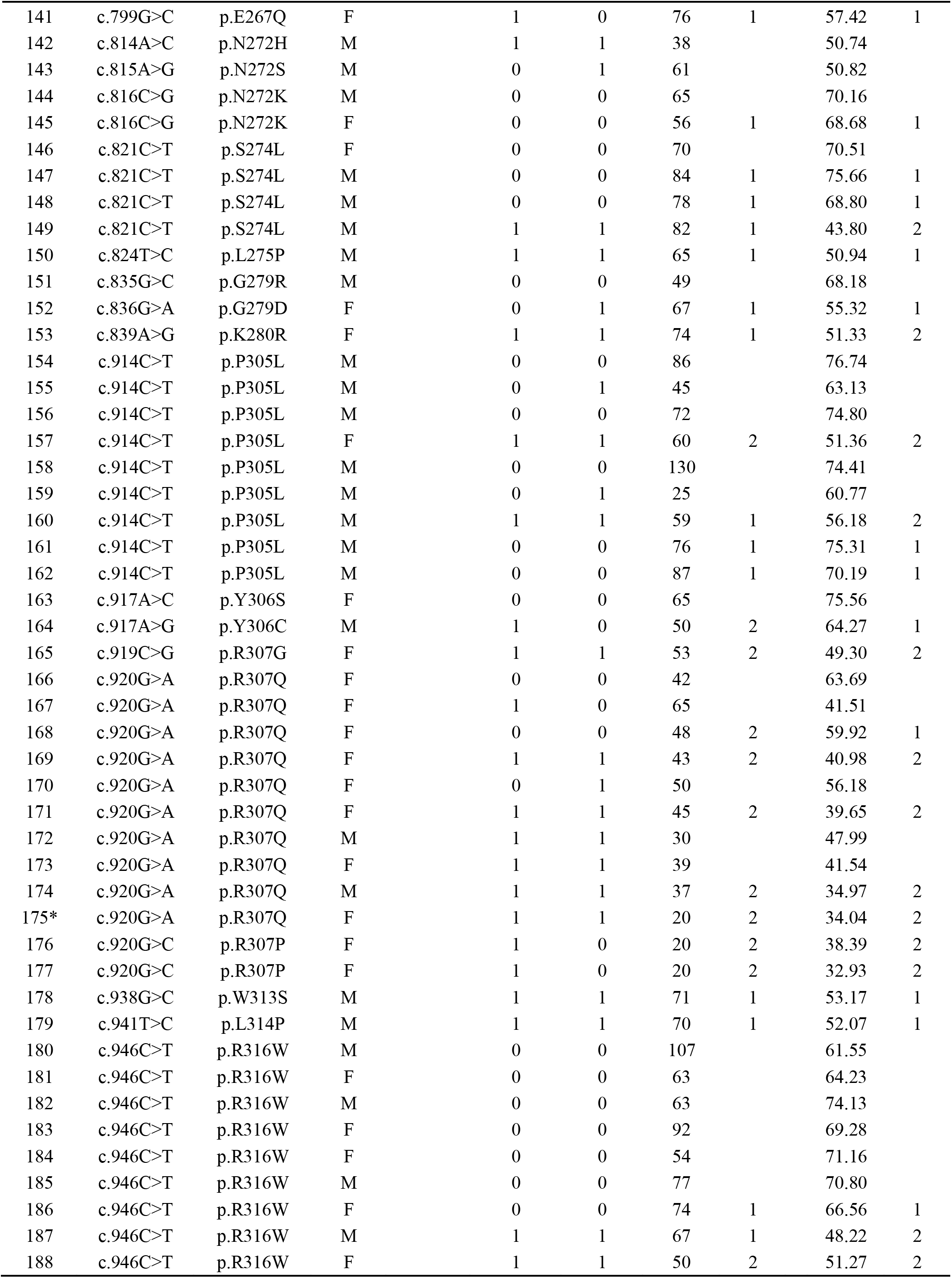
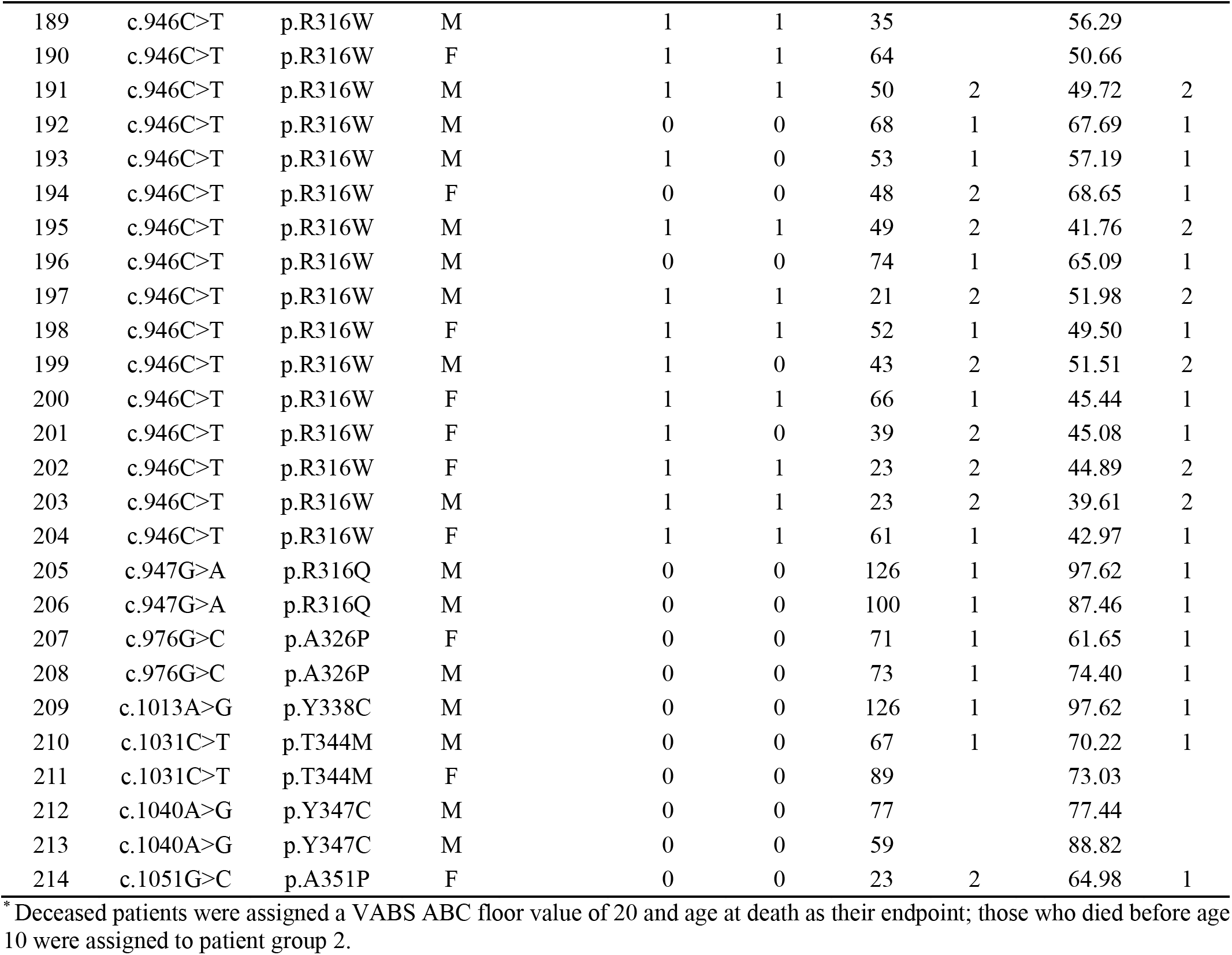
Patient-level classification and prediction results for all individuals with KIF1A pathogenic or likely pathogenic variants.

**Supplementary Table 5.** Ablation analysis in the prediction of most recent VABS ABC.

| Model | RMSE | MAE | $R^2$ | *Pearson r* |
| --- | --- | --- | --- | --- |
| ***Variant-level model*** |  |  |  |  |
| Full model | 18.254 | 14.545 | 0.195 | 0.444 |
| Remove molecular features | 19.117 | 15.813 | 0.117 | 0.376 |
| Remove computational scores | 19.628 | 15.111 | 0.070 | 0.343 |
| ***Patient-level model*** |  |  |  |  |
| Full model | 16.980 | 13.297 | 0.419 | 0.647 |
| Remove age | 17.317 | 13.896 | 0.395 | 0.630 |
| Remove seizures/EEG | 18.493 | 14.229 | 0.310 | 0.557 |
| Remove seizures/EEG, and age | 18.630 | 14.462 | 0.300 | 0.551 |
| Remove seizures/EEG and molecular features | 20.313 | 15.554 | 0.168 | 0.410 |
| Remove seizures/EEG and computational scores | 18.982 | 14.597 | 0.273 | 0.528 |
| Remove seizures/EEG, molecular features, and age | 20.695 | 15.987 | 0.136 | 0.374 |
| Remove seizures/EEG, computational scores, and age | 19.075 | 14.798 | 0.266 | 0.517 |

Computational scores include REVEL, gMVP, ESM, AlphaMissense, MisFit-D, and MisFit-S scores.
Molecular features include movement, diffusion, velocity, run length, dwell time, and conservation.
RMSE: Root mean square error, MAE: Mean absolute error, $R^2$ : Coefficient of determination.

**Supplementary Table 6.** Ablation analysis in the prediction of patient group.

| Model | Evaluation metrics |  |  |  |
| --- | --- | --- | --- | --- |
| | RMSE | MAE | $R^2$ | *Pearson r* |
| ***Variant-level model*** |  |  |  |  |
| Full model | 0.431 | 0.375 | 0.075 | 0.284 |
| Remove molecular features | 0.448 | 0.388 | 0.002 | 0.197 |
| Remove computational scores | 0.459 | 0.386 | -0.049 | 0.197 |
| ***Patient-level model*** |  |  |  |  |
|  | Accuracy | Sensitivity | Specificity | AUC |
| Full model | 0.809 | 0.869 | 0.712 | 0.834 |
| Remove age | 0.750 | 0.774 | 0.712 | 0.831 |
| Remove seizures/EEG | 0.794 | 0.869 | 0.673 | 0.804 |
| Remove seizures/EEG, and age | 0.728 | 0.821 | 0.577 | 0.789 |
| Remove seizures/EEG and molecular features | 0.706 | 0.798 | 0.558 | 0.753 |
| Remove seizures/EEG and computational scores | 0.735 | 0.810 | 0.615 | 0.732 |
| Remove seizures/EEG, molecular features, and age | 0.691 | 0.786 | 0.539 | 0.703 |
| Remove seizures/EEG, computational scores, and age | 0.632 | 0.726 | 0.481 | 0.662 |

Computational scores include REVEL, gMVP, ESM, AlphaMissense, MisFit-D, and MisFit-S scores.
Molecular features include movement, diffusion, velocity, run length, dwell time, and conservation.
RMSE: Root mean square error, MAE: Mean absolute error, $R^2$ : Coefficient of determination, AUC: Area under the curve.

